# Patterns of mental disorders in a nationwide child psychiatric sample (*N*=67,815): A DREAMS study

**DOI:** 10.1101/2024.02.29.24303557

**Authors:** Malindi van der Mheen, Josjan Zijlmans, Daniël van der Doelen, Helen Klip, Rikkert M. van der Lans, I. Hyun Ruisch, Ymkje Anna de Vries, Jacintha M. Tieskens, Marleen Wildschut, Jan K. Buitelaar, Pieter J. Hoekstra, Ramón J.L. Lindauer, Arne Popma, Robert R.J.M. Vermeiren, Wouter Staal, DREAMS consortium, Tinca J.C. Polderman

**Affiliations:** Amsterdam UMC, location University of Amsterdam, department of Child and Adolescent Psychiatry and Levvel, Academic Center for Child and Adolescent Psychiatry and Amsterdam Public Health Research Institute, Amsterdam UMC, all Amsterdam, the Netherlands; Amsterdam UMC, location Vrije Universiteit, department of Child and Adolescent Psychiatry & Psychosocial Care and Amsterdam Public Health Research Institute, Amsterdam UMC, both Amsterdam, the Netherlands; Levvel, Academic Center for Child and Adolescent Psychiatry, Amsterdam, the Netherlands; Karakter, Academic Center for Child and Adolescent Psychiatry, Nijmegen, the Netherlands; Department of Cognitive Neuroscience, Donders Institute for Brain, Cognition and Behaviour, Radboudumc, Nijmegen, The Netherlands; LUMC Curium - Child and Adolescent Psychiatry, Leiden UMC, Leiden, the Netherlands; University of Amsterdam, Research Institute of Child Development and Education (RICDE), Amsterdam, the Netherlands; Youz, Parnassia Group, The Hague, the Netherlands; Accare Child Study Center and University of Groningen, University Medical Center Groningen, Department of Child and Adolescent Psychiatry, both Groningen, the Netherlands; Levvel, Academic Center for Child and Adolescent Psychiatry, Amsterdam Public Health Research Institute, Amsterdam UMC, location Vrije Universiteit, department of Child and Adolescent Psychiatry & Psychosocial Care (all Amsterdam), Karakter, Academic Center for Child and Adolescent Psychiatry, Nijmegen, LUMC Curium - Child and Adolescent Psychiatry, Leiden UMC, Leiden, and Accare Child Study Center, Groningen, all the Netherlands

**Keywords:** Comorbidity, Prevalence, Diagnosis, Child Psychiatry, Adolescent Psychiatry

## Abstract

**Objective:** To provide a comprehensive overview of the prevalence and comorbidity patterns of mental disorders in a large, nationwide child and adolescent psychiatry sample.

**Methods:** We retrieved data on DSM diagnoses from medical records of children (0.5-23 years old) who received care at a DREAMS center between 2015 and 2019. DREAMS is a consortium of four academic centers for child and adolescent psychiatry in the Netherlands that provide both outpatient and inpatient care. Diagnoses were assigned in regular clinical practice.

**Results:** Between 2015 and 2019, 67,815 children received care at a DREAMS center (age at admission *M*=11.0 years, *SD=*4.3; 59.7% male). Of these children, 48,342 (71.3%) had a registered DSM disorder. The most prevalent primary diagnoses were ASD (34.1%), ADHD (24.4%) and trauma and stressor-related disorders (8.7%). Approximately half of all children (47.4%) had at least one comorbid diagnosis, of which intellectual disabilities were the most prevalent (14.0%).

**Conclusion:** Diagnostic patterns across sex and age as well as comorbidity patterns were generally consistent with previous research, but the prevalence of ASD and ADHD was higher than in other studies. Real-world diagnostic information such as presented here is essential to understand the use of DSM-5 in clinical practice, put differences between contexts and countries into perspective, and ultimately improve our diagnostic protocols and treatments.

## INTRODUCTION

Mental problems are a leading cause of disability in children and adolescents (hereafter referred to as children), associated with significant impairment on both the individual and societal level^1^. Comorbidity adds to this burden since the co-occurrence of multiple mental disorders has even more negative implications for quality of life, prognosis, and functional outcomes. For example, children with attention-deficit/hyperactivity disorder (ADHD) and a comorbid mental disorder are more likely to have a higher symptom severity, greater impairment, and poorer academic and behavioral outcomes (e.g., delinquency) than children with ADHD alone^2–4^. Similarly, comorbidity with an anxiety disorder has been associated with lower quality of life, a higher risk of recurrence, suicidality, longer duration of the disorder, more impairment, and decreased treatment response^5–8^. In other mental disorders, similar negative implications of comorbidity have been found (e.g., ^9,10–12^).

In addition, comorbidity can be relevant to what type of treatment is most appropriate^8,10,13,14^. Therefore, from a clinical perspective, gaining insight into comorbidity of mental disorders in children is crucial. Doing so is also fundamental from an epidemiological perspective to inform policies on general (nationwide) interventions, educational needs, protecting vulnerable groups, or clinical care. As most mental disorders emerge in childhood or adolescence^15,16^, understanding the epidemiology of comorbidity in children may also provide information on the etiology of mental disorders. Comorbidity patterns of mental disorders may be an indication of overlapping risk factors or shared etiology, and may as such also inform potential shared treatment options.

In the past decades, studies examining specific mental disorders have shown that comorbidity rates are high^17^. For example, in Autism Spectrum Disorder (ASD), reported prevalence rates of comorbidity of mental disorders in children vary from 55 to 94%, the most frequently reported comorbid diagnoses being ADHD, anxiety disorders, intellectual disabilities, and disruptive, impulse-control, and conduct disorders^18,19^. In ADHD, comorbidity rates are approximately 70%, mainly accounted for by disruptive, impulse-control, and conduct disorders, learning disorders, and anxiety and depressive disorders^20,21^. In other mental disorders, comorbidity also appears to be the rule rather than the exception. This seems to be more apparent in females than in males and rates increase with age^22^.

Although previous studies have examined comorbidity in specific mental disorders, studies providing a comprehensive overview of comorbid mental disorders in a large clinic-referred sample of children who receive psychiatric care are lacking. Furthermore, comorbidity patterns vary widely among studies and, generally, sample sizes are limited. Therefore, the current study aimed to provide a comprehensive overview of the prevalence and co-occurrence of mental disorders in a child and adolescent psychiatry sample that is large (*N*=67,815), covers a period of multiple years (i.e., 5 years), and represents large parts of the Netherlands. In addition, we aimed to examine differences in co-occurring mental disorders between age groups (i.e., 0-12 years old versus 13-23 years old) and between males and females.

## METHODS

### Study population

The current study used data from the Dutch REsearch in child and Adolescent Mental health (DREAMS) consortium (www.dreams-study.nl). DREAMS is a consortium of four academic centers for child and adolescent psychiatry in the Netherlands, i.e.: Accare, Levvel, LUMC Curium, and Karakter. These centers provide outpatient and inpatient care to children with varying mental health problems in the northern, western, and eastern part of the Netherlands. Of all children (0-20 years old) in the Netherlands, approximately 90% live in the catchment area of a DREAMS center, covering both urban and rural regions^23^. We used data from electronic health records of all children (aged 6 months through 23 years at time of admission) who received care at a DREAMS center between 2015 and 2019, and merged the summary statistics of each DREAMS center for this study.

As data were retrieved from medical records, this study was not subject to the Dutch Medical Research Involving Human Subjects Act, as confirmed by the Medical Ethical Committee of the Amsterdam UMC. Patients and their legal caregivers were informed that data collected within the framework of regular care could be used for scientific research.

### Diagnosis of mental disorders

#### DSM-IV-TR and DSM-5 diagnoses

Diagnoses of mental disorders were retrieved from electronic health records. Mental disorders were classified by mental health professionals according to DSM-IV-TR^24^ or DSM-5^25^ criteria following standard diagnostic procedures. That is, diagnoses were made in regular clinical practice following diagnostic procedures at the discretion of the mental health professional. In general, in 2015 and 2016, mental disorders were diagnosed according to DSM-IV-TR criteria. From 2017 through 2019, DSM-5 criteria were used. However, if a child only had a DSM-5 diagnosis in 2015 or 2016 or only had a DSM-IV-TR diagnosis in 2017 through 2019, we used the available diagnosis. For DSM-IV-TR, we included only axis I, II, and IV to exclude general medical conditions as well as the global functioning scale which does not exist in DSM-5.

#### Primary and comorbid diagnoses

We distinguished between primary and comorbid diagnoses. According to the DSM-5, the primary diagnosis concerns the mental health problems that are the main focus of clinical attention. We defined comorbid diagnoses as all non-primary diagnoses registered in a patient’s electronic health record between 2015 and 2019.

In general, mental health professionals explicitly defined in a child’s electronic health record which diagnosis was the primary one. The DREAMS centers use different electronic health record systems and one of the systems did not allow for differentiation between primary and comorbid diagnoses. In this case, if a child had more than one diagnosis, we considered the diagnosis listed first to be the primary one, as per the DSM-5 guidelines.

If more than one primary diagnosis was registered for a child, we selected the most recently registered primary diagnosis. We labeled the other diagnosis or diagnoses as comorbid. If two or more primary diagnoses were registered for a child on the exact same date, a computer algorithm randomly selected one of these diagnoses as primary. Again, we labeled the other diagnosis or diagnoses as comorbid.

#### Harmonizing DSM-IV-TR and DSM-5 diagnoses

To harmonize DSM-IV-TR and DSM-5 diagnoses, we categorized all diagnoses following DSM-5 diagnostic classes. First, all DSM-IV-TR and DSM-5 diagnoses were recoded into more generic DSM diagnoses to exclude subtypes and specifiers (see step 1 in Table S1, available online, for an example). Subtypes refer to mutually exclusive subgroups within a diagnosis (e.g., recurrent major depressive disorder versus single episode of a major depressive disorder). Specifiers are not mutually exclusive and provide more information on descriptive features of a diagnosis, such as onset and severity (e.g., early onset, severe, with atypical features).

Second, we recoded the diagnoses into broader DSM-5 diagnostic classes (see step 2 in Table S1**Error! Reference source not found.**, available online, for an example).

Third, we removed duplicate DSM-5 diagnostic classes for the same patient as it was unclear whether duplicates a) represented actual comorbidity (e.g., multiple specific phobias for the same patient), b) were due to changes in an individual’s mental health problems over time (e.g., major depressive disorder versus major depressive disorder in remission), c) were due to changes in a mental health professional’s diagnostic view (e.g., ADHD hyperactive-impulsive type versus ADHD combined type) or d) were due to registration errors.

Fourth, we constructed an overview of primary and five-year (i.e., 2015 through 2019) comorbid DSM-5 diagnostic classes. For privacy reasons, we only included primary DSM-5 diagnostic classes that occurred at least 50 times in our sample.

## RESULTS

### Sociodemographic characteristics

A flowchart of the study sample can be found in Figure 1**Error! Reference source not found.**. In total, 67,815 children received care at a DREAMS center between 2015 and 2019. Information on DSM disorders was available for 50,097 children (73.9%). Information on DSM disorders was missing due to lack of requirement of registration and administrative errors. Of these 50,097 children, 570 (1.1%) only had a registered V-code in their medical record (i.e., other conditions that may be a focus of clinical attention, such as relational or educational problems). Furthermore, in the medical records of 1,185 children (2.4%) it was registered that the DSM diagnosis had been delayed. Therefore, in total, 48,342 children (71.3% of *N =* 67,815) had a registered DSM disorder. Their mean age was 11.1 years at time of admission (range 0.5 to 23.5 years, *SD* = 4.3 years) and 61.3% were male. At time of admission, males’ mean age was 10.2 years (range 0.6 to 23.9 years, *SD =* 4.2 years) and females’ mean age was 12.5 years (range 0.5 to 23.8 years, *SD* = 4.2 years). Sociodemographic characteristics of the study sample are presented in Table 1.

**Figure 1.**
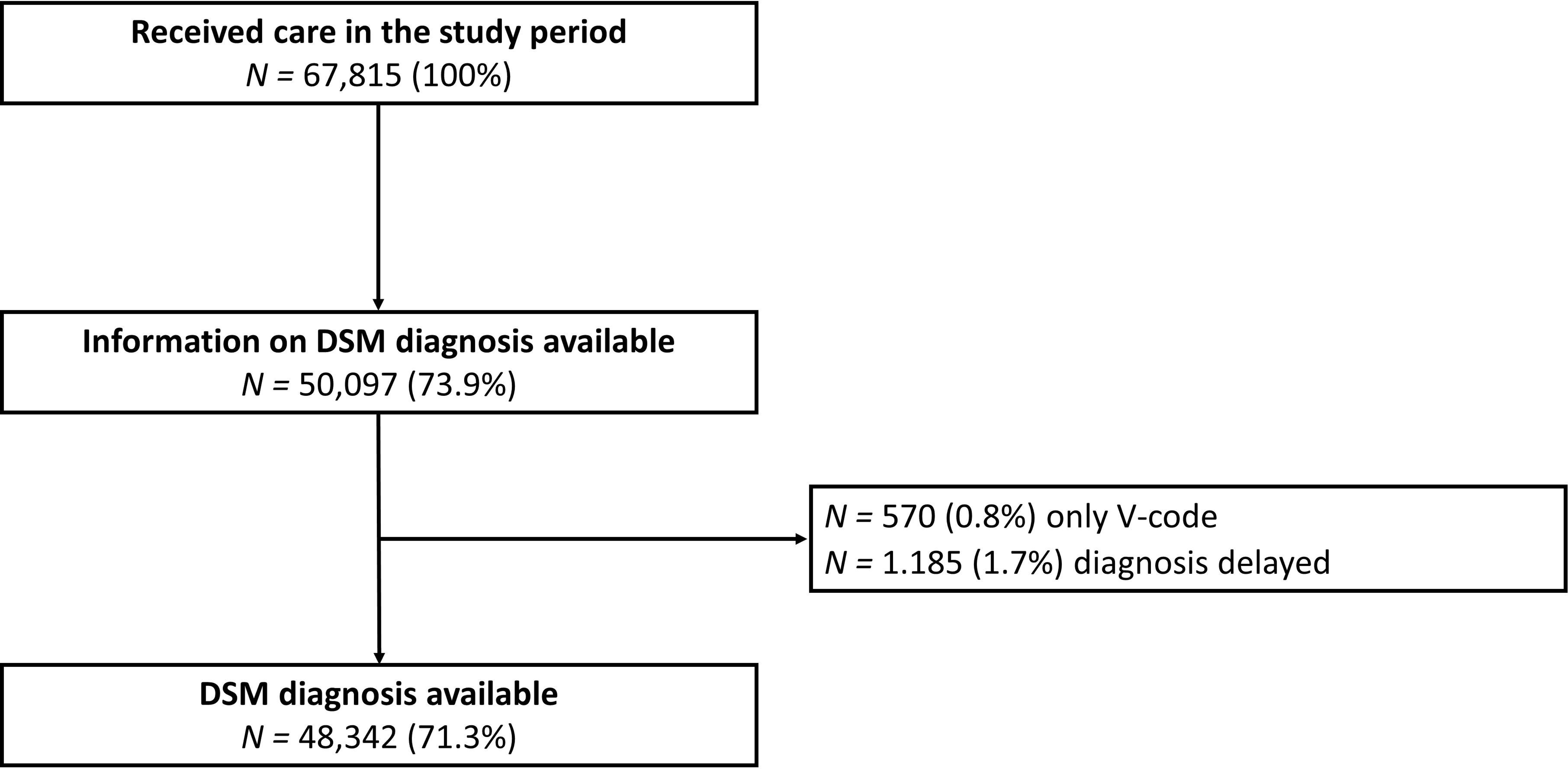
Study sample flowchart. Note. All reported percentages represent the proportion of the total population that received care in the study period 2015-2019 (i.e., *N* = 67,815).

**Table 1.**
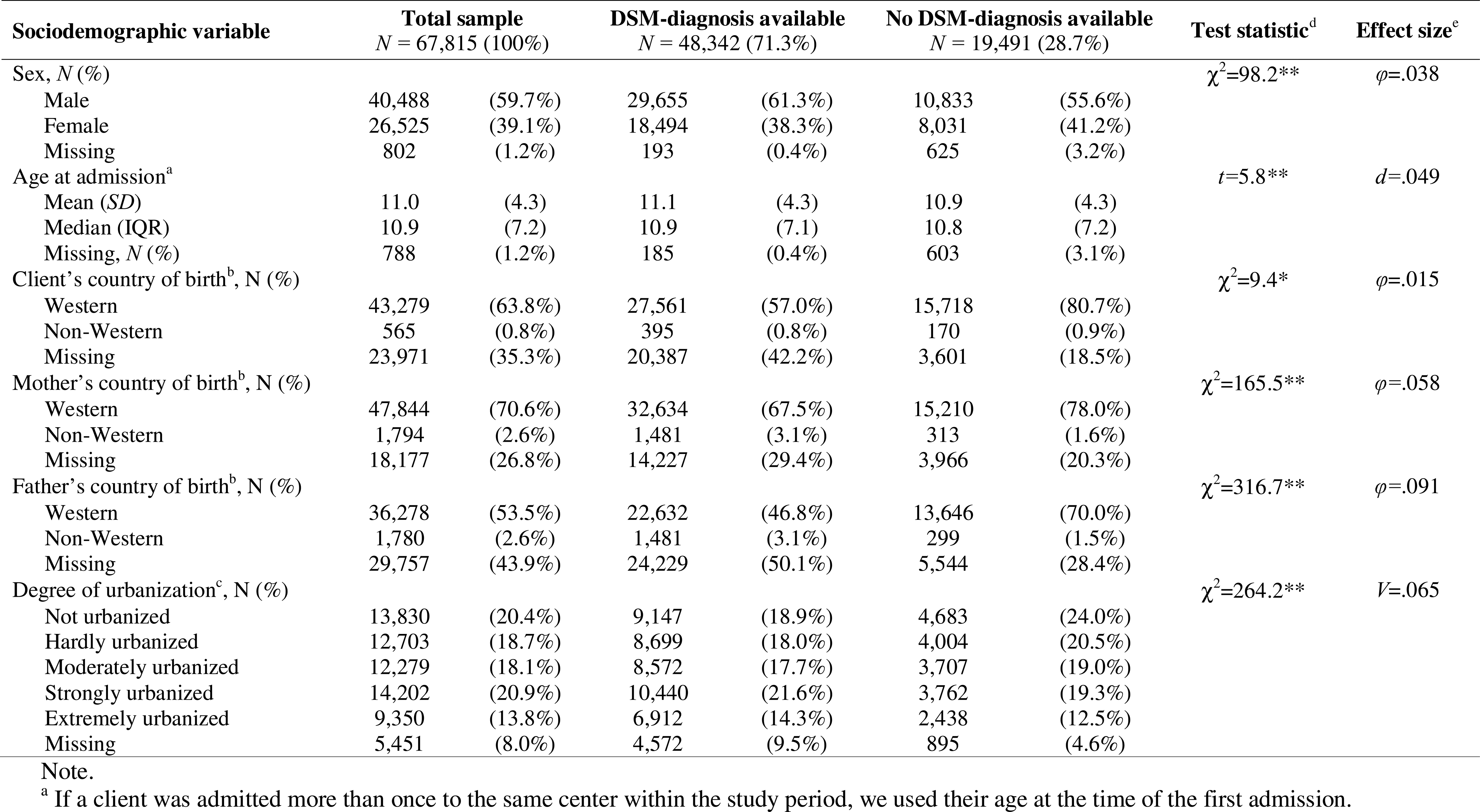
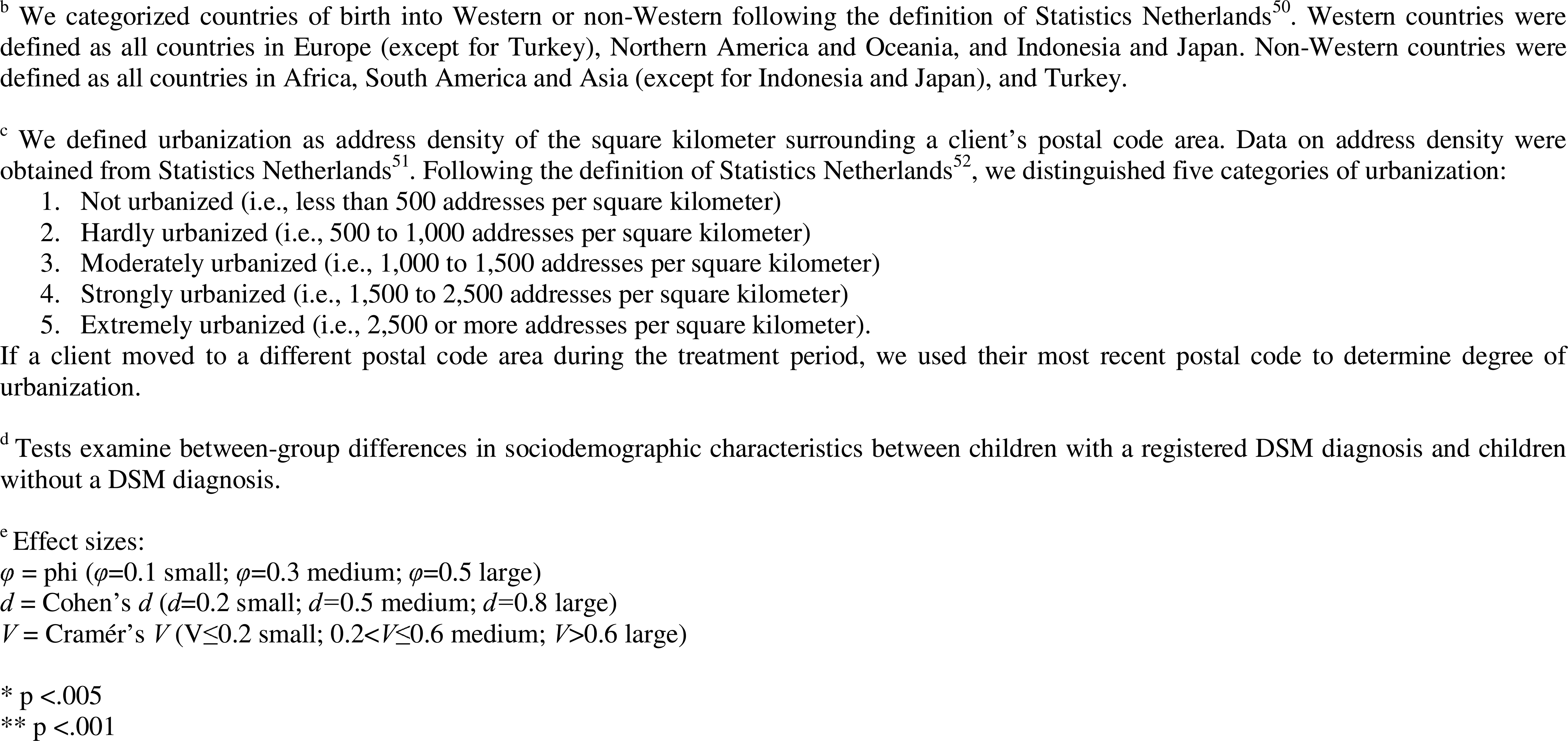
Sociodemographic characteristics of the study sample.

| Sociodemographic variable | Total sample<br>*N* = 67,815 (100%) |  | DSM-diagnosis available<br>*N* = 48,342 (71.3%) |  | No DSM-diagnosis available<br>*N* = 19,491 (28.7%) |  | Test statistic<sup>d</sup> | Effect size<sup>e</sup> |
| --- | --- | --- | --- | --- | --- | --- | --- | --- |
| Sex, *N* (%) | | | | | | | $\chi^2=98.2^{**}$ | $\varphi=.038$ |
| Male | 40,488 | (59.7%) | 29,655 | (61.3%) | 10,833 | (55.6%) |  |  |
| Female | 26,525 | (39.1%) | 18,494 | (38.3%) | 8,031 | (41.2%) |  |  |
| Missing | 802 | (1.2%) | 193 | (0.4%) | 625 | (3.2%) |  |  |
| Age at admission<sup>a</sup> | | | | | | | $t=5.8^{**}$ | $d=.049$ |
| Mean (*SD*) | 11.0 | (4.3) | 11.1 | (4.3) | 10.9 | (4.3) |  |  |
| Median (*IQR*) | 10.9 | (7.2) | 10.9 | (7.1) | 10.8 | (7.2) |  |  |
| Missing, *N* (%) | 788 | (1.2%) | 185 | (0.4%) | 603 | (3.1%) |  |  |
| Client's country of birth<sup>b</sup>, *N* (%) | | | | | | | $\chi^2=9.4^*$ | $\varphi=.015$ |
| Western | 43,279 | (63.8%) | 27,561 | (57.0%) | 15,718 | (80.7%) |  |  |
| Non-Western | 565 | (0.8%) | 395 | (0.8%) | 170 | (0.9%) |  |  |
| Missing | 23,971 | (35.3%) | 20,387 | (42.2%) | 3,601 | (18.5%) |  |  |
| Mother's country of birth<sup>b</sup>, *N* (%) | | | | | | | $\chi^2=165.5^{**}$ | $\varphi=.058$ |
| Western | 47,844 | (70.6%) | 32,634 | (67.5%) | 15,210 | (78.0%) |  |  |
| Non-Western | 1,794 | (2.6%) | 1,481 | (3.1%) | 313 | (1.6%) |  |  |
| Missing | 18,177 | (26.8%) | 14,227 | (29.4%) | 3,966 | (20.3%) |  |  |
| Father's country of birth<sup>b</sup>, *N* (%) | | | | | | | $\chi^2=316.7^{**}$ | $\varphi=.091$ |
| Western | 36,278 | (53.5%) | 22,632 | (46.8%) | 13,646 | (70.0%) |  |  |
| Non-Western | 1,780 | (2.6%) | 1,481 | (3.1%) | 299 | (1.5%) |  |  |
| Missing | 29,757 | (43.9%) | 24,229 | (50.1%) | 5,544 | (28.4%) |  |  |
| Degree of urbanization<sup>c</sup>, *N* (%) | | | | | | | $\chi^2=264.2^{**}$ | $V=.065$ |
| Not urbanized | 13,830 | (20.4%) | 9,147 | (18.9%) | 4,683 | (24.0%) |  |  |
| Hardly urbanized | 12,703 | (18.7%) | 8,699 | (18.0%) | 4,004 | (20.5%) |  |  |
| Moderately urbanized | 12,279 | (18.1%) | 8,572 | (17.7%) | 3,707 | (19.0%) |  |  |
| Strongly urbanized | 14,202 | (20.9%) | 10,440 | (21.6%) | 3,762 | (19.3%) |  |  |
| Extremely urbanized | 9,350 | (13.8%) | 6,912 | (14.3%) | 2,438 | (12.5%) |  |  |
| Missing | 5,451 | (8.0%) | 4,572 | (9.5%) | 895 | (4.6%) |  |  |

Note.
<sup>a</sup> If a client was admitted more than once to the same center within the study period, we used their age at the time of the first admission.
<sup>b</sup> We categorized countries of birth into Western or non-Western following the definition of Statistics Netherlands<sup>50</sup>. Western countries were defined as all countries in Europe (except for Turkey), Northern America and Oceania, and Indonesia and Japan. Non-Western countries were defined as all countries in Africa, South America and Asia (except for Indonesia and Japan), and Turkey.
<sup>c</sup> We defined urbanization as address density of the square kilometer surrounding a client's postal code area. Data on address density were obtained from Statistics Netherlands<sup>51</sup>. Following the definition of Statistics Netherlands<sup>52</sup>, we distinguished five categories of urbanization:
1. Not urbanized (i.e., less than 500 addresses per square kilometer) 2. Hardly urbanized (i.e., 500 to 1,000 addresses per square kilometer) 3. Moderately urbanized (i.e., 1,000 to 1,500 addresses per square kilometer) 4. Strongly urbanized (i.e., 1,500 to 2,500 addresses per square kilometer) 5. Extremely urbanized (i.e., 2,500 or more addresses per square kilometer).
If a client moved to a different postal code area during the treatment period, we used their most recent postal code to determine degree of urbanization.
<sup>d</sup> Tests examine between-group differences in sociodemographic characteristics between children with a registered DSM diagnosis and children without a DSM diagnosis.
<sup>e</sup> Effect sizes:
$\phi$ = phi ( $\phi=0.1$ small; $\phi=0.3$ medium; $\phi=0.5$ large)
$d$ = Cohen's $d$ ( $d=0.2$ small; $d=0.5$ medium; $d=0.8$ large)
$V$ = Cramér's $V$ ( $V \leq 0.2$ small; $0.2 < V \leq 0.6$ medium; $V > 0.6$ large)
\* $p < .005$
\*\* $p < .001$

To examine group differences in sociodemographic characteristics between children with a registered DSM diagnosis and children without a registered DSM diagnosis, we conducted a t-test for the continuous variable age and χ^2^ tests for categorical variables (i.e., sex, birth country, and degree of urbanization). As shown in Table 1, all group differences were statistically significant. However, the effect sizes indicated negligible differences between the groups.

### Primary and comorbid diagnoses

#### Total sample

Figure 2 shows the prevalence of primary and comorbid diagnoses in the total sample. The numbers behind Figure 2 can be found in Table S2, available online. The most prevalent primary diagnoses were ASD (34.1%) followed by ADHD (24.4%) and trauma and stressor-related disorders (8.7%). Primary diagnoses that occurred less than 50 times in our sample and that were not included in the results were: bipolar and related disorders, dissociative disorder, medication-induced movement disorder, neurocognitive disorder, paraphilic disorder, sexual dysfunction, and sleep-wake disorder.

**Figure 2.**
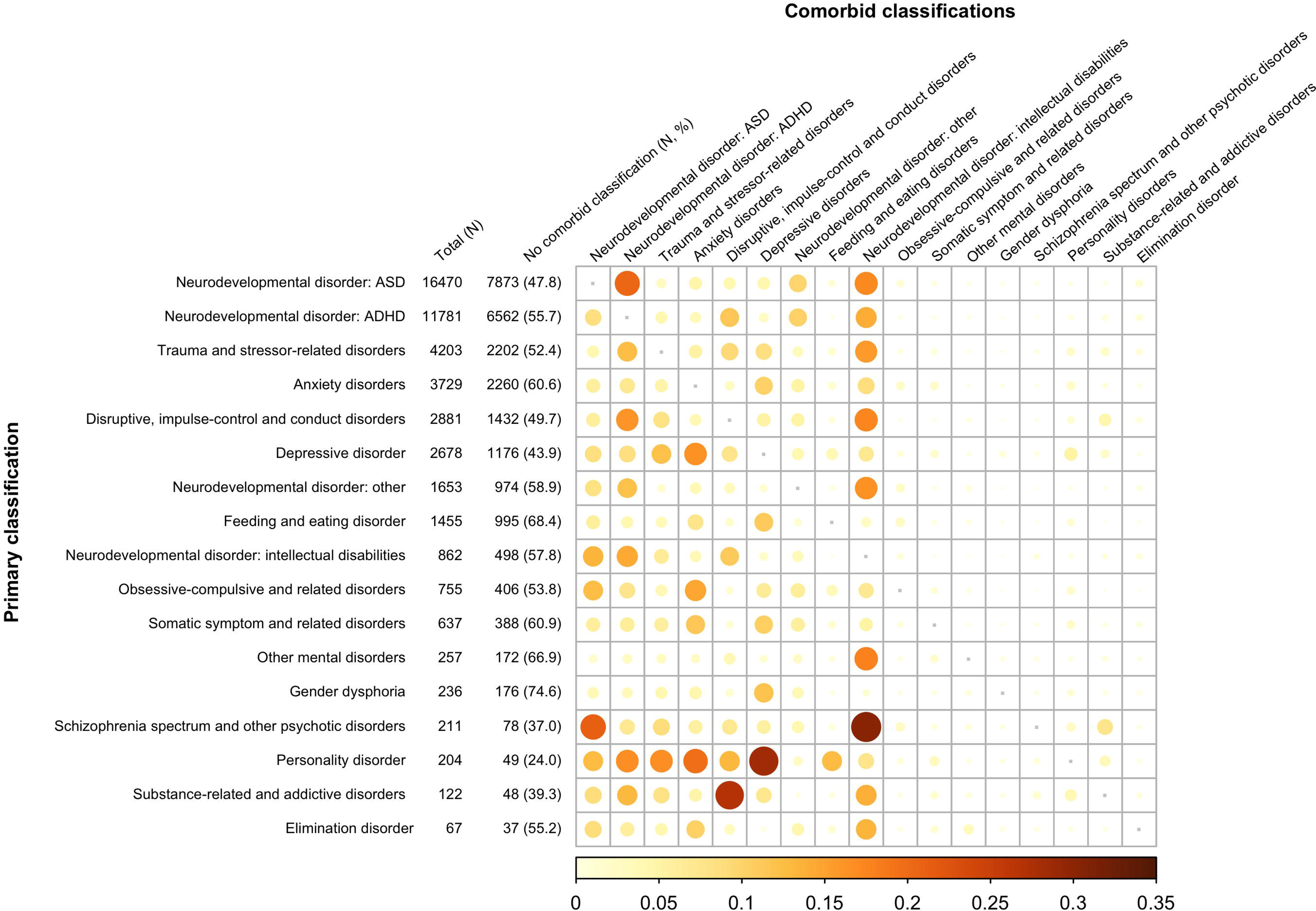
Primary and five-year comorbid DSM-5 diagnoses in the total child and adolescent psychiatric sample (N = 48,342). Note. Primary diagnoses are listed in order of prevalence. The size and color intensity of the circles correspond to the prevalence of a comorbidity for a particular primary diagnosis. Prevalence rates in the color key range from 0% to 35%. For privacy reasons, only primary diagnoses that occurred *N* > 50 are shown.

Approximately half of the children (47.4%) had one or more comorbid diagnoses. 32.3% of all children had one comorbid diagnosis, 11.5% had two, and 3.7% had three or more comorbid diagnoses. Furthermore, 28.0% had at least one V-code in addition to a mental disorder diagnosis (see Table S2, available online). Table S3, available online, provides a specification of the number of comorbid diagnoses per primary diagnosis.

Primary diagnoses of a personality disorder were most often accompanied by a comorbid diagnosis (see Figure 2). Only 24.0% of children with a personality disorder did not have any comorbid diagnosis. Depressive disorders were the most prevalent comorbid diagnosis in children with a personality disorder (28.4%). Of note, personality disorders were not often registered as a comorbid diagnosis for other primary diagnoses (*N* = 557; 1.2%) and were not very prevalent primary diagnoses (*N* = 204; 0.4%).

Overall, intellectual disabilities were the most prevalent comorbid diagnoses (14.0%). Intellectual disabilities were particularly prevalent as comorbid diagnoses in children with a primary diagnosis of a schizophrenia spectrum and other psychotic disorder (30.3%). Also, a primary diagnosis of substance-related and addictive disorders with comorbid disruptive, impulse-control and conduct disorders was relatively prevalent (27.0%).

Of note, comorbidity patterns were not symmetrical. For example, a comorbid diagnosis of ADHD in children with a primary diagnosis of ASD was more prevalent (20.6%) than comorbid ASD in children with ADHD (8.4%).

#### Sex specific results

Figure 3 shows the prevalence of primary and comorbid diagnoses for males and females separately. The numbers behind Figure 3 can be found in Tables S4 and S5, available online. In males (*N =* 29,655), the most prevalent primary diagnoses were ASD (41.0%), followed by ADHD (28.8%), disruptive, impulse-control and conduct disorders (6.3%), and trauma and stressor-related disorders (6.2%). In females (*N =* 18,494), the most prevalent primary diagnoses were ASD (23.2%), followed by ADHD (17.0%), trauma and stressor-related disorders (12.6%), and anxiety disorders (12.5%).

**Figure 3.**
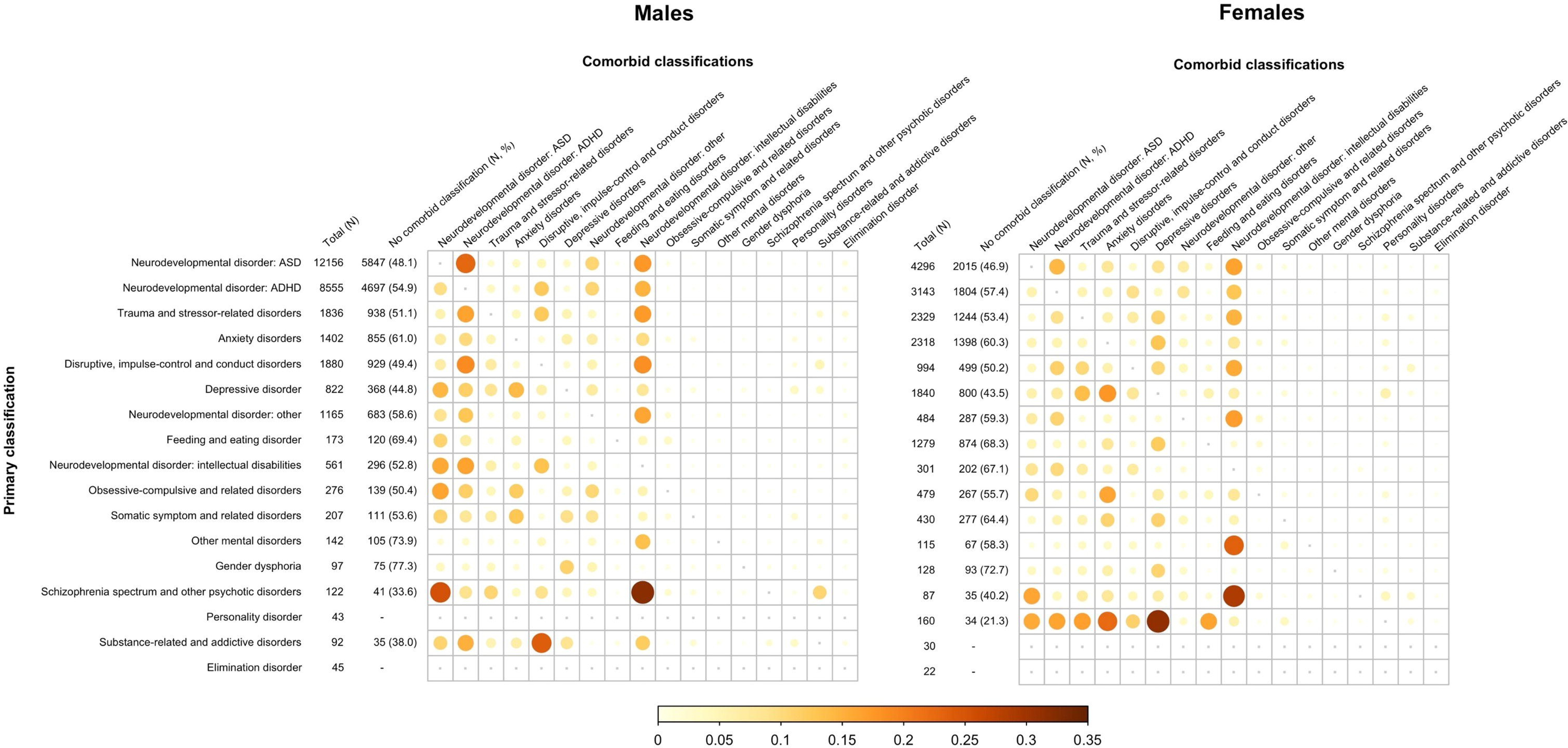
Primary and five year comorbid DSM-5 diagnoses by sex (males N = 29,655; and females N = 18,494). Note. The size and color intensity of the circles correspond to the prevalence of a comorbidity for a particular primary diagnosis. Prevalence rates in the color key range from 0% to 35%. For privacy reasons, only primary diagnoses that occurred N > 50 in the overall sample are listed. Data on prevalence rates of comorbidities are only presented for primary diagnoses that occurred N > 50 in the subsample.

Though ASD and ADHD were the most prevalent primary diagnoses in both males and females, there were differences in prevalence patterns of primary diagnoses. First, in males, ASD and ADHD accounted for 69.8% of primary diagnoses. In females, ASD and ADHD accounted for a smaller proportion of primary diagnoses, namely 40.2%. Second, several primary diagnoses were more prevalent in females than in males. For example, trauma and stressor-related disorders accounted for 12.6% of primary diagnoses in females and for 6.2% in males. The same was true for anxiety disorders (females 12.5% versus males 4.7%), depressive disorders (females 9.9% versus males 2.8%) and feeding and eating disorders (females 6.9% versus males 0.6%).

As to comorbidity, 48.3% of males and 46.2% of females had at least one comorbid diagnosis. In females, primary diagnoses of a personality disorder were often accompanied by a comorbid diagnosis (78.7%). In males, these findings were not shown as fewer than 50 males had a primary diagnosis of a personality disorder. In both males and females, intellectual disabilities were prevalent comorbid diagnoses (males 15.4%, females 11.8%). Intellectual disabilities were particularly prevalent in children with a primary diagnosis of schizophrenia spectrum and other psychotic disorders (males 32.0%, females 28.7%). In males, comorbid diagnoses of ASD and ADHD were more prevalent than in females. That is, 13.6% of males (*N* = 4,023) had a comorbid ADHD diagnosis versus 7.8% of females (*N =* 1,439) and 5.4% of males (*N* = 1,602) had a comorbid ASD diagnosis versus 4.1% of females (*N* = 765).

#### Age specific results

Figure 4 shows the prevalence of primary and comorbid diagnoses by age at time of diagnosis. The numbers behind Figure 4 can be found in Tables S6 and S7, available online. In children who were 0 through 12 years old at time of diagnosis (*N* = 15,646), the most prevalent primary diagnoses were ASD (39.7%), ADHD (29.9%), and trauma and stressor-related disorders (7.8%). In children who were 13 through 23 years old at time of diagnosis (*N* = 32,513), ASD (31.5%), ADHD (21.6%), and trauma and stressor-related disorders (9.0%) were the most prevalent primary diagnoses as well, although ASD and ADHD accounted for a smaller proportion of primary classifications in older children. Another noticeable difference was that 0.8% of 0 through 12-year-olds had a primary diagnosis of a depressive disorder versus 7.8% of 13 through 23-year-olds. Moreover, several diagnoses, including schizophrenia spectrum and other psychotic disorders, personality disorders, and substance-related and addictive disorders, were nearly entirely absent in 0 through 12-year-olds.

**Figure 4.**
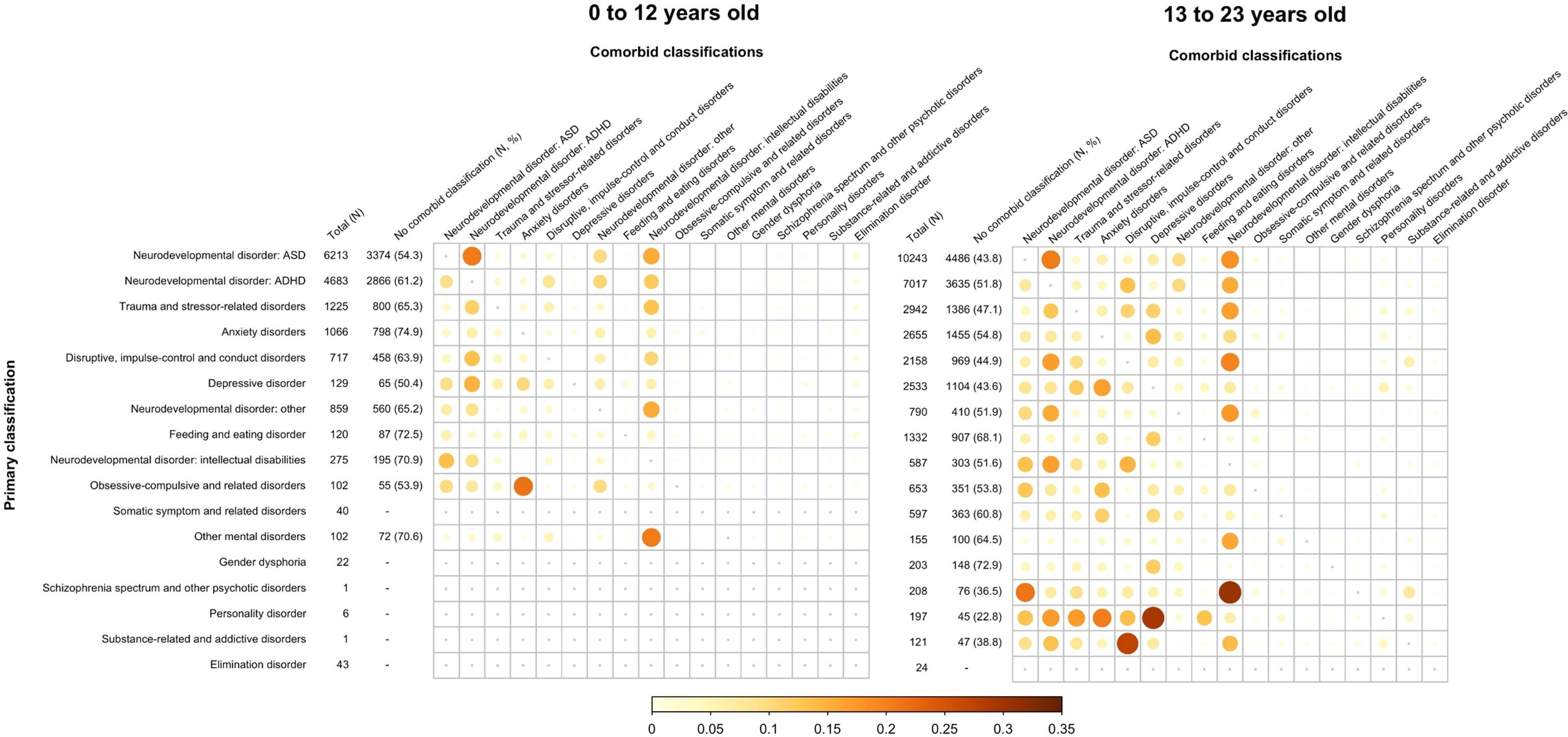
Primary and five-year comorbid DSM-5 diagnoses by age at time of primary diagnosis (0 through 12 years old N = 15,646; 13 through 23 years old N = 32,513). Note. The size and color intensity of the circles correspond to the prevalence of a comorbidity for a particular primary diagnosis. Prevalence rates in the color key range from 0% to 35%. For privacy reasons, only primary diagnoses that occurred N > 50 in the overall sample are listed. Data on prevalence rates of comorbidities are only presented for primary diagnoses that occurred N > 50 in the subsample.

Of all 0 through 12-year-olds, 39.7% had at least one comorbid diagnosis. In 13 through 23-year-olds this percentage was higher, namely 51.3%. Primary diagnoses of a personality disorder were most often accompanied by a comorbid diagnosis in older children (77.2%), particularly with a comorbid depressive disorder (29.4%). In younger children, these data were not shown as fewer than 50 young children had a primary diagnosis of a personality disorder. In both age groups, intellectual disabilities were the most prevalent comorbid diagnosis (younger children 13.0%, older children 14.5%). In older children, intellectual disabilities were particularly comorbid with a primary diagnosis of a schizophrenia spectrum and other psychotic disorder (30.3%). Another common specific comorbidity in older children was a primary diagnosis of a substance-related and addictive disorder with a comorbid disruptive, impulse-control and conduct disorder (27.3%). Data on schizophrenia spectrum and other psychotic disorders and substance-related and addictive disorders were not shown for younger children (*N* < 50). In younger children, the most common specific comorbidities were a primary diagnosis of obsessive compulsive disorder (OCD) with a comorbid anxiety disorder (21.6% versus 13.5% in older children), ASD with comorbid ADHD (with 20.7% versus 20.6% similar to older children) and other mental disorders (e.g., other specified mental disorder due to another medical condition) with a comorbid intellectual disability (20.6% versus 16.1% in older children).

Figures S1 and S2 and Tables S8 through S11, available online, show the prevalence of primary and comorbid DSM-5 diagnoses by sex and age at time of diagnosis.

## DISCUSSION

The aim of the current study was to provide a comprehensive overview of the prevalence of 5-year primary and comorbid mental disorder diagnoses in a large sample of children who received psychiatric care. In the total sample, ASD and ADHD were the most prevalent primary diagnoses, accounting for 58.5% (ASD 34.1% and ADHD 24.4%) of the population. Approximately half of all children had at least one comorbid diagnosis. We observed substantial differences between males and females, both in terms of primary diagnoses and comorbid diagnoses. 69.8% of males had a primary diagnosis of ASD or ADHD versus 40.2% of females. In females, primary diagnoses of trauma and stressor-related disorders, anxiety disorders, depressive disorders, and feeding and eating disorders were more prevalent than in males, consistent with previous reports^26–29^. Also, though a similar proportion of males and females had at least one comorbid diagnosis (males 48.3%, females 46.2%), ASD and ADHD were more often registered as a comorbid diagnosis in males (19.0%) than in females (11.9%), which is in line with literature as well^30^. Regarding age at time of diagnosis, primary diagnoses of ASD and ADHD were more prevalent in children aged 0-12 (69.6%) than in children aged 13-23 (53.1%) and comorbidity was more prevalent in children aged 13-23 (51.3%) than in children aged 0-12 (39.7%). These differences between age groups are also consistent with literature^22,31,32^.

Regarding primary diagnoses, previous studies generally have found substantially lower prevalence rates of ASD and ADHD in youth referred for psychiatric care^10^. First, this difference may be due to the fact that, in the Netherlands, children with ASD or ADHD are often referred to centers for child and adolescent psychiatry, whereas these children are more often treated by a pediatrician in other countries^33^. Second, this difference may reflect a discrepancy in prevalence rates between countries. In a different sample of children referred for psychiatric care in the Netherlands (*N* = 1,402), Jansen and colleagues^34^ also found relatively high prevalence rates for ASD (35.1%) and ADHD (33.6%). Our findings are in line with the observation that population-based prevalence rates of ASD differ widely across geographical areas, which may be due to socio-cultural and socio-economic determinants^35^. Third, this difference may be explained by methodological differences between studies. That is, previous studies conducted semi-structured clinical interviews to assess diagnostic criteria, whereas the current study used data on diagnoses determined by mental health professionals following standard diagnostic procedures in clinical practice.

Although prevalence rates of comorbidity of mental disorders vary widely in literature, the prevalence of comorbidity seemed to be lower in the current study than generally found in previous studies^17,36,37^. Previous research has shown that mental health professionals tend to overlook comorbid mental disorders^38,39^, which may have resulted in a lower prevalence of comorbidity in the current study compared with studies that used standardized structured assessment methods. Furthermore, the current study only reported on comorbidity between diagnostic classes (e.g., an anxiety disorder with a depressive disorder), which will have lowered our rates of comorbidity compared to studies that include all types of comorbidity (e.g., an anxiety disorder with another anxiety disorder). In the current study, it was not possible to include all types of comorbidity as it was not clear whether comorbidity within the same diagnostic class represented actual comorbidity or noise in diagnostic or registration procedures. These explanations may also account for the fact that we found comparable overall rates of comorbidity in males and females, whereas previous studies observed more comorbidity in females^40^.

In line with previous research, patterns of comorbidity were not symmetrical in our sample^41,42^, suggesting that some classifications are more likely to be seen as primary, while others are more likely to be seen as secondary to other problems. For example, ASD with comorbid ADHD (20.6%) was more prevalent than ADHD with comorbid ASD (8.4%). Of note, the DSM-5 is the first edition allowing mental health professionals to classify comorbid ADHD in individuals with ASD and vice versa. Previous research has also found an asymmetrical comorbidity pattern regarding ASD and ADHD.

Some results regarding comorbidity might rather be explained by the diagnostic system than by actual diagnostic symptoms. For instance, a primary diagnosis of a personality disorder was most often accompanied by a comorbid diagnosis, whereas intellectual disorders were most often registered as comorbid diagnoses. This may be due to the multiaxial system of the DSM-IV-TR. In this system, personality disorders and intellectual disabilities were classified on Axis II. Doing so required a diagnosis of at least one mental health disorder on Axis I, which may have resulted in these high comorbidity rates.

Contrary to what the nosology of the DSM suggests^25^, the comorbidity rates found in the current study indicate that diagnostic classes are not discrete entities. The separation and dichotomization of psychopathology in the DSM has been frequently critiqued^43–46^. Furthermore, it has frequently been argued that the DSM categorizes psychopathology into too many distinct diagnostic classes^45–47^, which our results seem to confirm considering the high comorbidity rates. Additionally, the DSM is based on science and clinical knowledge originating from psychiatry, neurology, and epidemiology, but has limited etiological validity^48^. This is relevant, as previous studies have shown that comorbidity of mental disorders results from shared genetic and environmental risk factors^45^. Alternative diagnosis frameworks for psychopathology that may overcome these shortcomings are the Hierarchical Taxonomy of Psychopathology (HiTOP^47,48^) and the Research Domain Criteria (RDoC^46,49^). HiTOP is a dimensional diagnosis system which is based on state-of-the-art scientific evidence. In contrast to the dichotomous view of the DSM, HiTOP considers psychopathology to be a continuum. Moreover, HiTOP allows mental health professionals to classify psychopathology at multiple levels. This may offer a solution for the issues regarding comorbidity and high variability in the presentation of DSM-diagnoses. Similarly to HiTOP, RDoC conceptualizes mental disorders on a continuum. However, in contrast to the DSM and HiTOP, the RDoC framework considers the etiology of mental disorders^46,49^. Although both frameworks also come with disadvantages^45^, it would be useful to further examine the clinical and scientific value of these frameworks.

The current study has several strengths. We provided a comprehensive overview of primary and comorbid mental disorders in a substantial sample of children who received psychiatric care in The Netherlands over a 5-year period. We included data from four academic centers for child and adolescent psychiatry representing large parts of the Netherlands, which enhances the generalizability of our results. Furthermore, all diagnoses were determined by mental health professionals, enhancing ecological validity. However, it should also be noted that diagnostic procedures differ between clinical centers and mental health professionals. Another limitation is that the DREAMS centers use different electronic health record systems, which complicated harmonizing the data. More specifically, in one DREAMS center, the electronic health record system did not allow for mental health professionals to explicitly define which diagnosis was the focus of clinical attention. Therefore, as per DSM-5 guidelines, if a child had more than one registered DSM diagnosis, we considered the diagnosis listed first to be the primary one. Finally, the data were retrieved from electronic health records which may have included registration errors and were not always unambiguously interpretable. For this reason, we did not present comorbidity patterns within the same diagnostic class (e.g., specific phobia with comorbid panic disorder).

In sum, we provided a comprehensive overview of the prevalence of primary and 5-year comorbid mental disorders in a large sample of children who received psychiatric care. Diagnostic patterns were generally in line with previous research, but the prevalence of ASD and ADHD was higher than in other studies. Real world diagnostic information such as presented here is essential to understand the use of DSM-5 in clinical practice, put differences between contexts and countries into perspective, and ultimately improve our diagnostic protocols and treatments.

## Data Availability

All data produced in the present study are available upon reasonable request to the authors

## Author Contributions

Conceptualization: Van der Mheen, Zijlmans, Polderman, Klip, Staal, Van der Doelen, Van der Lans, Ruisch, Tieskens, Buitelaar, Hoekstra, Lindauer, Popma, Vermeiren

Data curation: Van der Mheen, Zijlmans, Van der Doelen, Ruisch, Van der Lans

Formal Analysis: Van der Mheen, Zijlmans

Investigation: Van der Mheen, Zijlmans, Polderman

Methodology: Van der Mheen, Zijlmans, Polderman Supervision: Polderman

Visualization: De Vries

Writing – original draft: Van der Mheen

Writing – review & editing: Van der Mheen, Zijlmans, Polderman, Staal, Van der Doelen, Klip, Van der Lans, Ruisch, De Vries, Tieskens, Wildschut, Buitelaar, Hoekstra, Lindauer, Popma, Vermeiren, DREAMS consortium

## Acknowledgements

Funding: This study was funded by the Association of Netherlands Municipalities. The funder had no role in study design, data collection, analysis, or interpretation nor in writing the paper.

