## Supplemental table 1 for "Patterns of mental disorders in a nationwide child psychiatric sample (*N*=67,815): A DREAMS study"

Table S 1. Example of how DSM-diagnoses were harmonized into DSM-5 diagnostic classes.

| Raw data<br>Diagnosis with subtype and specifiers | Step 1<br>Diagnosis without subtype and specifiers | Step 2<br>DSM-5 diagnostic class |
|---|---|---|
| • Major depressive disorder, single episode, mild<br>• Major depressive disorder, recurrent episode, severe with psychotic features | Major depressive disorder | Depressive disorders |
| • Persistent depressive disorder (dysthymic disorder), early onset<br>• Persistent depressive disorder (dysthymic disorder), with atypical features | Persistent depressive disorder | |
