## Supplemental table 2 for "Patterns of mental disorders in a nationwide child psychiatric sample (*N*=67,815): A DREAMS study"

Table S 2. Prevalence of primary and five-year comorbid DSM-5 diagnoses in the total child and adolescent psychiatric sample ($N$ = 48,342).

| | | | | | Comorbid diagnosis | | | | | | | | | | | | | | | | |
|---|---|---|---|---|---|---|---|---|---|---|---|---|---|---|---|---|---|---|---|---|---|
| | Total | | No comorbid diagnosis | V-code | Neurodevelopmental disorder: ASD | Neurodevelopmental disorder: ADHD | Trauma and stressor-related disorders | Anxiety disorders | Disruptive, impulse-control and conduct disorders | Depressive disorder | Neurodevelopmental disorder: other | Feeding and eating disorder | Neurodevelopmental disorder: intellectual disabilities | Obsessive-compulsive and related disorders | Somatic symptom and related disorders | Other mental disorders | Gender dysphoria | Schizophrenia spectrum and other psychotic disorders | Personality disorder | Substance-related and addictive disorders | Elimination disorder |
| **Primary diagnosis** | n | % | % | % | % | % | % | % | % | % | % | % | % | % | % | % | % | % | % | % | % |
| Neurodevelopmental disorder: ASD | 16,470 | 34.1% | 47.8% | 25.3% | - | 20.6% | 2.8% | 4.8% | 4.3% | 4.6% | 9.8% | 1.3% | 17.3% | 1.3% | 0.6% | 0.3% | 0.2% | 0.4% | 0.4% | 0.4% | 1.4% |
| Neurodevelopmental disorder: ADHD | 11,781 | 24.4% | 55.7% | 26.9% | 8.4% | - | 4.2% | 3.6% | 11.4% | 2.5% | 10.2% | 0.3% | 14.1% | 0.4% | 0.3% | 0.3% | 0.1% | 0.1% | 0.6% | 0.8% | 1.2% |
| Trauma and stressor-related disorders | 4,203 | 8.7% | 52.4% | 33.8% | 4.3% | 12.5% | - | 5.3% | 9.4% | 8.4% | 3.2% | 1.6% | 15.7% | 0.6% | 0.9% | 0.3% | 0.2% | 0.4% | 2.0% | 2.1% | 1.1% |
| Anxiety disorders | 3,729 | 7.7% | 60.6% | 25.6% | 5.9% | 7.2% | 5.0% | - | 2.5% | 10.2% | 5.2% | 1.7% | 8.8% | 2.0% | 2.0% | 0.2% | 0.3% | 0.2% | 2.2% | 0.7% | 0.5% |
| Disruptive, impulse-control and conduct disorders | 2,881 | 6.0% | 49.7% | 41.2% | 5.9% | 16.2% | 8.2% | 4.0% | - | 5.3% | 5.1% | 1.0% | 17.4% | 0.2% | 0.2% | 0.6% | 0.1% | 0.2% | 1.5% | 4.6% | 0.9% |
| Depressive disorder | 2,678 | 5.5% | 43.9% | 41.0% | 8.5% | 8.7% | 12.2% | 16.5% | 7.6% | - | 4.2% | 4.1% | 7.0% | 1.0% | 1.9% | 0.4% | 1.0% | 0.5% | 5.2% | 2.1% | 0.4% |
| Neurodevelopmental disorder: other | 1,653 | 3.4% | 58.9% | 24.0% | 8.3% | 12.2% | 1.8% | 3.8% | 3.3% | 1.6% | - | 0.5% | 16.5% | 2.2% | 0.4% | 0.9% | 0.1% | 0.1% | 0.3% | 0.2% | 0.8% |
| Feeding and eating disorder | 1,455 | 3.0% | 68.4% | 14.4% | 5.8% | 4.2% | 3.6% | 7.6% | 1.9% | 11.1% | 1.5% | - | 2.7% | 2.4% | 0.5% | 0.6% | 0.1% | 0.1% | 1.5% | 0.1% | 0.2% |
| Neurodevelopmental disorder: intellectual disabilities | 862 | 1.8% | 57.8% | 25.8% | 13.3% | 14.4% | 6.6% | 3.7% | 11.1% | 2.0% | 3.6% | 0.1% | - | 1.0% | 0.1% | 0.1% | 0.1% | 0.8% | 0% | 0.7% | 0.9% |
| Obsessive-compulsive and related disorders | 755 | 1.6% | 53.8% | 20.1% | 12.5% | 7.8% | 4.0% | 14.6% | 1.5% | 6.6% | 6.4% | 3.6% | 7.0% | - | 1.1% | 0.5% | 0.1% | 0.1% | 1.9% | 0.1% | 0.1% |
| Somatic symptom and related disorders | 637 | 1.3% | 60.9% | 21.5% | 6.0% | 6.3% | 6.1% | 11.5% | 1.4% | 10.5% | 5.8% | 1.4% | 5.2% | 0.8% | - | 0.3% | 0.2% | 0.3% | 2.0% | 0% | 0.8% |
| Other mental disorders | 257 | 0.5% | 66.9% | 16.0% | 1.9% | 2.7% | 2.7% | 2.3% | 3.9% | 1.6% | 2.3% | 0.8% | 17.9% | 0% | 1.9% | - | 0% | 0% | 0% | 0.8% | 0.4% |
| Gender dysphoria | 236 | 0.5% | 74.6% | 45.3% | 3.8% | 3.4% | 4.2% | 4.7% | 1.7% | 11.9% | 3.8% | 0.4% | 1.3% | 0.4% | 0.4% | 0.8% | - | 0.4% | 1.3% | 0% | 0% |
| Schizophrenia spectrum and other psychotic disorders | 211 | 0.4% | 37.0% | 19.0% | 21.3% | 7.1% | 9.0% | 5.7% | 7.1% | 5.7% | 4.3% | 0.9% | 30.3% | 2.4% | 0.9% | 0.9% | 0.5% | - | 2.4% | 7.6% | 0% |
| Personality disorder | 204 | 0.4% | 24.0% | 51.5% | 12.7% | 16.7% | 16.7% | 19.6% | 13.2% | 28.4% | 2.5% | 12.7% | 7.8% | 1.0% | 2.9% | 0% | 0.5% | 1.0% | - | 3.4% | 0% |
| Substance-related and addictive disorders | 122 | 0.3% | 39.3% | 41.8% | 9.0% | 13.1% | 8.2% | 4.9% | 27.0% | 7.4% | 0% | 0.8% | 13.9% | 0% | 1.6% | 0.8% | 0% | 1.6% | 4.1% | - | 0.8% |
| Elimination disorder | 67 | 0.1% | 55.2% | 40.3% | 9.0% | 6.0% | 4.5% | 10.4% | 3.0% | 0% | 4.5% | 1.5% | 13.4% | 0% | 1.5% | 3.0% | 0% | 0% | 0% | 0% | - |
