## Supplemental table 3 for "Patterns of mental disorders in a nationwide child psychiatric sample (*N*=67,815): A DREAMS study"

Table S 3. Number of comorbid DSM diagnoses per primary diagnosis.

| | | Number of comorbid DSM diagnoses | | | | | | | |
|---|---|---|---|---|---|---|---|---|---|
| | | 0 | | 1 | | 2 | | 3 or more | |
| Primary DSM diagnosis | Total *N* | *N* | % | *N* | % | *N* | % | *N* | % |
| Neurodevelopmental disorder: ASD | 16,470 | 7,868 | 47.8% | 5,966 | 36.2% | 2,047 | 12.4% | 589 | 3.6% |
| Neurodevelopmental disorder: ADHD | 11,781 | 6,567 | 55.7% | 3,685 | 31.3% | 1,201 | 10.2% | 328 | 2.8% |
| Trauma and stressor-related disorders | 4,203 | 2,204 | 52.4% | 1,283 | 30.5% | 526 | 12.5% | 190 | 4.5% |
| Anxiety disorders | 3,729 | 2,260 | 60.6% | 1,000 | 26.8% | 354 | 9.5% | 115 | 3.1% |
| Disruptive, impulse-control and conduct disorders | 2,881 | 1,432 | 49.7% | 950 | 33.0% | 365 | 12.7% | 134 | 4.7% |
| Depressive disorder | 2,678 | 1,176 | 43.9% | 958 | 35.8% | 369 | 13.8% | 175 | 6.5% |
| Neurodevelopmental disorder: other | 1,653 | 973 | 58.9% | 500 | 30.2% | 144 | 8.7% | 36 | 2.2% |
| Feeding and eating disorder | 1,455 | 995 | 68.4% | 318 | 21.9% | 104 | 7.1% | 38 | 2.6% |
| Neurodevelopmental disorder: intellectual disabilities | 862 | 498 | 57.8% | 247 | 28.7% | 97 | 11.3% | 20 | 2.3% |
| Obsessive-compulsive and related disorders | 755 | 406 | 53.8% | 219 | 29.0% | 87 | 11.5% | 43 | 5.7% |
| Somatic symptom and related disorders | 637 | 388 | 60.9% | 155 | 24.3% | 60 | 9.4% | 34 | 5.3% |
| Other mental disorders | 257 | 172 | 66.9% | 72 | 28.0% | 11 | 4.3% | 2 | 0.8% |
| Gender dysphoria | 236 | 176 | 74.6% | 38 | 16.1% | 15 | 6.4% | 7 | 3.0% |
| Schizophrenia spectrum and other psychotic disorders | 211 | 78 | 37.0% | 66 | 31.3% | 43 | 20.4% | 24 | 11.4% |
| Personality disorder | 204 | 49 | 24.0% | 63 | 30.9% | 62 | 30.4% | 30 | 14.7% |
| Substance-related and addictive disorders | 122 | 48 | 39.3% | 43 | 35.2% | 21 | 17.2% | 10 | 8.2% |
| Elimination disorder | 67 | 37 | 55.2% | 21 | 31.3% | 8 | 11.9% | 1 | 1.5% |
| Overall | 48,342 | 25,405 | 52.6% | 15,615 | 32.3% | 5,536 | 11.5% | 1,786 | 3.7% |
