## Supplemental table 4 for "Patterns of mental disorders in a nationwide child psychiatric sample (*N*=67,815): A DREAMS study"

Table S 4. Primary and five year comorbid DSM-5 diagnoses in males (*N* = 29,655).

| | Total | | No comorbid diagnosis | V-code | **Comorbid diagnosis** | | | | | | | | | | | | | | | | |
|---|---|---|---|---|---|---|---|---|---|---|---|---|---|---|---|---|---|---|---|---|---|
| | | | | | Neurodevelopmental disorder: ASD | Neurodevelopmental disorder: ADHD | Trauma and stressor-related disorders | Anxiety disorders | Disruptive, impulse-control and conduct disorders | Depressive disorder | Neurodevelopmental disorder: other | Feeding and eating disorder | Neurodevelopmental disorder: intellectual disabilities | Obsessive-compulsive and related disorders | Somatic symptom and related disorders | Other mental disorders | Gender dysphoria | Schizophrenia spectrum and other psychotic disorders | Personality disorder | Substance-related and addictive disorders | Elimination disorder |
| **Primary diagnosis** | n | % | % | % | % | % | % | % | % | % | % | % | % | % | % | % | % | % | % | % | % |
| Neurodevelopmental disorder: ASD | 12,156 | 41.0% | 48.1% | 23.9% | - | 22.8% | 2.5% | 3.6% | 4.6% | 3.2% | 10.7% | 0.7% | 17.6% | 0.9% | 0.4% | 0.3% | 0.1% | 0.3% | 0.2% | 0.5% | 1.5% |
| Neurodevelopmental disorder: ADHD | 8,555 | 28.8% | 54.9% | 25.9% | 9.5% | - | 3.8% | 3.0% | 12.3% | 2.0% | 10.8% | 0.1% | 14.7% | 0.4% | 0.2% | 0.2% | 0.0% | 0.1% | 0.3% | 1.0% | 1.3% |
| Trauma and stressor-related disorders | 1,836 | 6.2% | 51.1% | 32.4% | 5.6% | 16.6% | - | 4.2% | 12.4% | 5.3% | 3.8% | 0.3% | 17.2% | 0.4% | 0.1% | 0.3% | 0.1% | 0.3% | 0.7% | 2.5% | 2.0% |
| Anxiety disorders | 1,402 | 4.7% | 61.0% | 25.8% | 7.1% | 10.0% | 4.9% | - | 2.4% | 6.1% | 6.7% | 0.9% | 9.8% | 1.5% | 1.5% | 0.1% | 0.4% | 0.2% | 1.5% | 1.2% | 0.9% |
| Disruptive, impulse-control and conduct disorders | 1,880 | 6.3% | 49.4% | 37.6% | 6.8% | 18.8% | 7.0% | 2.9% | - | 2.7% | 5.5% | 0.3% | 18.5% | 0.1% | 0.1% | 0.4% | 0.2% | 0.2% | 1.3% | 5.0% | 1.1% |
| Depressive disorder | 822 | 2.8% | 44.8% | 40.4% | 14.4% | 11.7% | 8.5% | 14.1% | 6.2% | - | 7.2% | 0.7% | 8.2% | 1.2% | 1.7% | 0.1% | 1.2% | 0.6% | 3.8% | 3.3% | 0.6% |
| Neurodevelopmental disorder: other | 1,165 | 3.9% | 58.6% | 24.0% | 8.8% | 12.7% | 1.5% | 3.6% | 3.6% | 1.4% | - | 0.3% | 16.2% | 2.0% | 0.3% | 0.9% | 0.0% | 0.1% | 0.1% | 0.3% | 1.0% |
| Feeding and eating disorder | 173 | 0.6% | 69.4% | 11.6% | 11.0% | 6.9% | 1.7% | 5.2% | 0.0% | 4.6% | 1.7% | - | 4.6% | 3.5% | 0.6% | 0.6% | 0.0% | 0.0% | 0.0% | 0.0% | 1.2% |
| Neurodevelopmental disorder: intellectual disabilities | 561 | 1.9% | 52.8% | 25.1% | 15.9% | 16.6% | 6.4% | 3.0% | 13.2% | 2.0% | 4.5% | 0.2% | - | 0.7% | 0.0% | 0.0% | 0.2% | 0.5% | 0.0% | 0.9% | 1.2% |
| Obsessive-compulsive and related disorders | 276 | 0.9% | 50.4% | 14.9% | 16.3% | 11.6% | 4.7% | 12.0% | 1.4% | 5.4% | 10.5% | 1.8% | 5.4% | - | 0.7% | 0.4% | 0.4% | 0.4% | 0.7% | 0.4% | 0.4% |
| Somatic symptom and related disorders | 207 | 0.7% | 53.6% | 21.3% | 11.1% | 8.2% | 7.2% | 12.6% | 1.9% | 9.2% | 8.7% | 0.5% | 4.8% | 1.9% | - | 0.0% | 0.0% | 0.0% | 2.4% | 0.0% | 1.9% |
| Other mental disorders | 142 | 0.5% | 73.9% | 14.1% | 2.1% | 2.8% | 2.1% | 1.4% | 4.2% | 0.7% | 3.5% | 0.7% | 13.4% | 0.0% | 0.7% | - | 0.0% | 0.0% | 0.0% | 1.4% | 0.7% |
| Gender dysphoria | 97 | 0.3% | 77.3% | 48.5% | 4.1% | 3.1% | 4.1% | 2.1% | 1.0% | 11.3% | 5.2% | 1.0% | 1.0% | 1.0% | 1.0% | 1.0% | - | 1.0% | 1.0% | 0.0% | 0.0% |
| Schizophrenia spectrum and other psychotic disorders | 122 | 0.4% | 33.6% | 16.4% | 25.4% | 9.0% | 10.7% | 4.1% | 9.0% | 4.9% | 4.1% | 0.0% | 32.0% | 2.5% | 1.6% | 0.8% | 0.8% | - | 0.8% | 10.7% | 0.0% |
| Personality disorder | 43 | 0.1% | - | - | - | - | - | - | - | - | - | - | - | - | - | - | - | - | - | - | - |
| Substance-related and addictive disorders | 92 | 0.3% | 38.0% | 42.4% | 10.9% | 15.2% | 5.4% | 6.5% | 23.9% | 8.7% | 0.0% | 1.1% | 12.0% | 0.0% | 2.2% | 0.0% | 0.0% | 2.2% | 3.3% | - | 1.1% |
| Elimination disorder | 45 | 0.2% | - | - | - | - | - | - | - | - | - | - | - | - | - | - | - | - | - | - | - |
