## Supplemental table 5 for "Patterns of mental disorders in a nationwide child psychiatric sample (*N*=67,815): A DREAMS study"

Table S 5. Primary and five year comorbid DSM-5 diagnoses in females ($N$ = 18,494).

| | Total | | | | Comorbid diagnosis | | | | | | | | | | | | | | | | |
|---|---|---|---|---|---|---|---|---|---|---|---|---|---|---|---|---|---|---|---|---|---|
| | | | No comorbid diagnosis | V-code | Neurodevelopmental disorder: ASD | Neurodevelopmental disorder: ADHD | Trauma and stressor-related disorders | Anxiety disorders | Disruptive, impulse-control and conduct disorders | Depressive disorder | Neurodevelopmental disorder: other | Feeding and eating disorder | Neurodevelopmental disorder: intellectual disabilities | Obsessive-compulsive and related disorders | Somatic symptom and related disorders | Other mental disorders | Gender dysphoria | Schizophrenia spectrum and other psychotic disorders | Personality disorder | Substance-related and addictive disorders | Elimination disorder |
| **Primary diagnosis** | n | % | % | % | % | % | % | % | % | % | % | % | % | % | % | % | % | % | % | % | % |
| Neurodevelopmental disorder: ASD | 4,296 | 23.2% | 46.9% | 29.1% | - | 14.4% | 3.8% | 8.1% | 3.2% | 8.8% | 7.1% | 3.1% | 16.6% | 2.2% | 1.3% | 0.4% | 0.4% | 0.4% | 1.0% | 0.2% | 1.0% |
| Neurodevelopmental disorder: ADHD | 3,143 | 17.0% | 57.4% | 29.6% | 5.7% | - | 5.3% | 5.3% | 9.1% | 4.0% | 8.4% | 0.9% | 12.9% | 0.6% | 0.7% | 0.4% | 0.1% | 0.1% | 1.5% | 0.4% | 1.0% |
| Trauma and stressor-related disorders | 2,329 | 12.6% | 53.4% | 34.6% | 3.2% | 9.2% | - | 6.1% | 7.0% | 10.9% | 2.6% | 2.7% | 14.7% | 0.8% | 1.5% | 0.3% | 0.3% | 0.5% | 3.0% | 1.9% | 0.4% |
| Anxiety disorders | 2,318 | 12.5% | 60.3% | 25.5% | 5.2% | 5.5% | 5.0% | - | 2.5% | 12.7% | 4.4% | 2.2% | 8.2% | 2.3% | 2.4% | 0.2% | 0.3% | 0.2% | 2.6% | 0.3% | 0.2% |
| Disruptive, impulse-control and conduct disorders | 994 | 5.4% | 50.2% | 47.9% | 4.2% | 11.4% | 10.4% | 5.9% | - | 10.5% | 4.4% | 2.4% | 15.6% | 0.6% | 0.3% | 0.8% | 0.1% | 0.3% | 1.8% | 3.8% | 0.5% |
| Depressive disorder | 1,840 | 9.9% | 43.5% | 41.1% | 5.9% | 7.4% | 13.7% | 17.7% | 8.2% | - | 2.9% | 5.7% | 6.5% | 0.9% | 2.0% | 0.5% | 0.9% | 0.5% | 5.8% | 1.5% | 0.3% |
| Neurodevelopmental disorder: other | 484 | 2.6% | 59.3% | 23.6% | 7.0% | 11.0% | 2.3% | 4.1% | 2.5% | 2.3% | - | 1.0% | 17.1% | 2.9% | 0.6% | 1.0% | 0.2% | 0.0% | 0.8% | 0.2% | 0.2% |
| Feeding and eating disorder | 1,279 | 6.9% | 68.3% | 14.7% | 5.1% | 3.8% | 3.8% | 7.9% | 2.1% | 12.0% | 1.5% | - | 2.4% | 2.3% | 0.5% | 0.6% | 0.1% | 0.1% | 1.7% | 0.1% | 0.1% |
| Neurodevelopmental disorder: intellectual disabilities | 301 | 1.6% | 67.1% | 26.9% | 8.6% | 10.3% | 7.0% | 5.0% | 7.3% | 2.0% | 2.0% | 0.0% | - | 1.7% | 0.3% | 0.3% | 0.0% | 1.3% | 0.0% | 0.3% | 0.3% |
| Obsessive-compulsive and related disorders | 479 | 2.6% | 55.7% | 23.2% | 10.2% | 5.6% | 3.5% | 16.1% | 1.5% | 7.3% | 4.0% | 4.6% | 7.9% | - | 1.3% | 0.6% | 0.0% | 0.0% | 2.5% | 0.0% | 0.0% |
| Somatic symptom and related disorders | 430 | 2.3% | 64.4% | 21.6% | 3.5% | 5.3% | 5.6% | 10.9% | 1.2% | 11.2% | 4.4% | 1.9% | 5.3% | 0.2% | - | 0.5% | 0.2% | 0.5% | 1.9% | 0.0% | 0.2% |
| Other mental disorders | 115 | 0.6% | 58.3% | 18.3% | 1.7% | 2.6% | 3.5% | 3.5% | 3.5% | 2.6% | 0.9% | 0.9% | 23.5% | 0.0% | 3.5% | - | 0.0% | 0.0% | 0.0% | 0.0% | 0.0% |
| Gender dysphoria | 128 | 0.7% | 72.7% | 42.2% | 3.9% | 3.9% | 4.7% | 6.3% | 2.3% | 10.9% | 3.1% | 0.0% | 1.6% | 0.0% | 0.0% | 0.8% | - | 0.0% | 1.6% | 0.0% | 0.0% |
| Schizophrenia spectrum and other psychotic disorders | 87 | 0.5% | 40.2% | 21.8% | 16.1% | 4.6% | 6.9% | 8.0% | 4.6% | 6.9% | 4.6% | 2.3% | 28.7% | 2.3% | 0.0% | 1.1% | 0.0% | - | 4.6% | 3.4% | 0.0% |
| Personality disorder | 160 | 0.9% | 21.3% | 56.3% | 15.6% | 16.3% | 16.9% | 22.5% | 11.3% | 31.3% | 3.1% | 16.3% | 5.6% | 0.6% | 3.8% | 0.0% | 0.6% | 0.6% | - | 1.9% | 0.0% |
| Substance-related and addictive disorders | 30 | 0,2% | - | - | - | - | - | - | - | - | - | - | - | - | - | - | - | - | - | - | - |
| Elimination disorder | 22 | 0,1% | - | - | - | - | - | - | - | - | - | - | - | - | - | - | - | - | - | - | - |
