## Supplemental table 6 for "Patterns of mental disorders in a nationwide child psychiatric sample (*N*=67,815): A DREAMS study"

Table S 6. Primary and five-year comorbid DSM-5 diagnoses by age at time of primary diagnosis: 0 through 12 years old ($N$ = 15,646).

| | Total | | | | Comorbid diagnosis | | | | | | | | | | | | | | | | |
|---|---|---|---|---|---|---|---|---|---|---|---|---|---|---|---|---|---|---|---|---|---|
| | | | No comorbid diagnosis | V-code | Neurodevelopmental disorder: ASD | Neurodevelopmental disorder: ADHD | Trauma and stressor-related disorders | Anxiety disorders | Disruptive. impulse-control and conduct disorders | Depressive disorder | Neurodevelopmental disorder: other | Feeding and eating disorder | Neurodevelopmental disorder: intellectual disabilities | Obsessive-compulsive and related disorders | Somatic symptom and related disorders | Other mental disorders | Gender dysphoria | Schizophrenia spectrum and other psychotic disorders | Personality disorder | Substance-related and addictive disorders | Elimination disorder |
| **Primary diagnosis** | n | % | % | % | % | % | % | % | % | % | % | % | % | % | % | % | % | % | % | % | % |
| Neurodevelopmental disorder: ASD | 6,213 | 39.7% | 54.3% | 24.1% | - | 20.7% | 1.6% | 2.6% | 3.2% | 0.8% | 9.9% | 0.8% | 15.7% | 0.5% | 0.2% | 0.2% | 0.1% | 0.0% | 0.0% | 0.0% | 2.0% |
| Neurodevelopmental disorder: ADHD | 4,683 | 29.9% | 61.2% | 30.6% | 9.4% | - | 2.8% | 2.6% | 8.6% | 0.7% | 10.5% | 0.2% | 12.4% | 0.3% | 0.1% | 0.4% | 0.0% | 0.0% | 0.0% | 0.0% | 1.5% |
| Trauma and stressor-related disorders | 1,225 | 7.8% | 65.3% | 35.6% | 3.5% | 11.6% | - | 2.9% | 5.9% | 1.1% | 3.5% | 0.5% | 13.0% | 0.1% | 0.0% | 0.2% | 0.1% | 0.0% | 0.0% | 0.1% | 1.9% |
| Anxiety disorders | 1,066 | 6.8% | 74.9% | 23.4% | 3.6% | 6.2% | 3.0% | - | 2.0% | 1.4% | 5.9% | 0.3% | 5.9% | 1.1% | 1.3% | 0.3% | 0.4% | 0.0% | 0.3% | 0.0% | 1.0% |
| Disruptive, impulse-control and conduct disorders | 717 | 4.6% | 63.9% | 39.2% | 4.6% | 13.4% | 4.3% | 3.6% | - | 0.4% | 5.9% | 0.3% | 10.7% | 0.0% | 0.0% | 0.6% | 0.1% | 0.0% | 0.0% | 0.0% | 1.3% |
| Depressive disorder | 129 | 0.8% | 50.4% | 55.0% | 9.3% | 14.7% | 6.2% | 10.1% | 6.2% | - | 6.2% | 2.3% | 6.2% | 0.0% | 0.0% | 0.8% | 0.0% | 0.0% | 1.6% | 0.0% | 1.6% |
| Neurodevelopmental disorder: other | 859 | 5.5% | 65.2% | 25.8% | 6.6% | 9.0% | 0.8% | 3.0% | 2.9% | 0.5% | - | 0.6% | 15.6% | 0.7% | 0.1% | 0.8% | 0.0% | 0.0% | 0.1% | 0.0% | 0.8% |
| Feeding and eating disorder | 120 | 0.8% | 72.5% | 12.5% | 5.8% | 3.3% | 3.3% | 4.2% | 3.3% | 1.7% | 3.3% | - | 4.2% | 0.8% | 0.8% | 0.0% | 0.0% | 0.0% | 0.8% | 0.0% | 1.7% |
| Neurodevelopmental disorder: intellectual disabilities | 275 | 1.8% | 70.9% | 22.2% | 13.5% | 9.5% | 2.5% | 0.4% | 3.3% | 0.0% | 3.3% | 0.4% | - | 0.0% | 0.0% | 0.0% | 0.0% | 0.0% | 0.0% | 0.0% | 0.7% |
| Obsessive-compulsive and related disorders | 102 | 0.7% | 53.9% | 15.7% | 9.8% | 7.8% | 3.9% | 21.6% | 1.0% | 1.0% | 9.8% | 1.0% | 2.9% | - | 0.0% | 1.0% | 0.0% | 0.0% | 1.0% | 0.0% | 0.0% |
| Somatic symptom and related disorders | 40 | 0.3% | - | - | - | - | - | - | - | - | - | - | - | - | - | - | - | - | - | - | - |
| Other mental disorders | 102 | 0.7% | 70.6% | 15.7% | 2.0% | 2.9% | 3.9% | 1.0% | 4.9% | 0.0% | 2.0% | 0.0% | 20.6% | 0.0% | 0.0% | - | 0.0% | 0.0% | 0.0% | 0.0% | 0.0% |
| Gender dysphoria | 22 | 0.1% | - | - | - | - | - | - | - | - | - | - | - | - | - | - | - | - | - | - | - |
| Schizophrenia spectrum and other psychotic disorders | 1 | 0.0% | - | - | - | - | - | - | - | - | - | - | - | - | - | - | - | - | - | - | - |
| Personality disorder | 6 | 0.0% | - | - | - | - | - | - | - | - | - | - | - | - | - | - | - | - | - | - | - |
| Substance-related and addictive disorders | 1 | 0.0% | - | - | - | - | - | - | - | - | - | - | - | - | - | - | - | - | - | - | - |
| Elimination disorder | 43 | 0.3% | - | - | - | - | - | - | - | - | - | - | - | - | - | - | - | - | - | - | - |
