## Supplemental table 7 for "Patterns of mental disorders in a nationwide child psychiatric sample (*N*=67,815): A DREAMS study"

Table S 7. Primary and five-year comorbid DSM-5 diagnoses by age at time of primary diagnosis: 13 through 23 years old (*N* = 32,513).

| | Total | | No comorbid diagnosis | V-code | Comorbid diagnosis | | | | | | | | | | | | | | | | |
|---|---|---|---|---|---|---|---|---|---|---|---|---|---|---|---|---|---|---|---|---|---|
| | | | | | Neurodevelopmental disorder: ASD | Neurodevelopmental disorder: ADHD | Trauma and stressor-related disorders | Anxiety disorders | Disruptive. impulse-control and conduct disorders | Depressive disorder | Neurodevelopmental disorder: other | Feeding and eating disorder | Neurodevelopmental disorder: intellectual disabilities | Obsessive-compulsive and related disorders | Somatic symptom and related disorders | Other mental disorders | Gender dysphoria | Schizophrenia spectrum and other psychotic disorders | Personality disorder | Substance-related and addictive disorders | Elimination disorder |
| **Primary diagnosis** | n | % | % | % | % | % | % | % | % | % | % | % | % | % | % | % | % | % | % | % | % |
| Neurodevelopmental disorder: ASD | 10,243 | 31.5% | 43.8% | 26.0% | - | 20.6% | 3.6% | 6.1% | 4.9% | 6.9% | 9.7% | 1.6% | 18.3% | 1.7% | 0.9% | 0.3% | 0.3% | 0.6% | 0.7% | 0.6% | 1.0% |
| Neurodevelopmental disorder: ADHD | 7,017 | 21.6% | 51.8% | 24.5% | 7.8% | - | 5.1% | 4.2% | 13.4% | 3.7% | 10.0% | 0.4% | 15.5% | 0.5% | 0.4% | 0.2% | 0.1% | 0.1% | 0.9% | 1.4% | 1.1% |
| Trauma and stressor-related disorders | 2,942 | 9.0% | 47.1% | 32.8% | 4.6% | 12.8% | - | 6.3% | 10.9% | 11.5% | 3.0% | 2.1% | 16.9% | 0.8% | 1.2% | 0.3% | 0.2% | 0.5% | 2.8% | 3.0% | 0.8% |
| Anxiety disorders | 2,655 | 8.2% | 54.8% | 26.5% | 6.9% | 7.6% | 5.8% | - | 2.7% | 13.7% | 5.0% | 2.3% | 9.9% | 2.3% | 2.3% | 0.1% | 0.3% | 0.3% | 3.0% | 0.9% | 0.2% |
| Disruptive, impulse-control and conduct disorders | 2,158 | 6.6% | 44.9% | 41.8% | 6.3% | 17.1% | 9.5% | 4.0% | - | 7.0% | 4.9% | 1.3% | 19.7% | 0.3% | 0.2% | 0.5% | 0.1% | 0.3% | 1.9% | 6.1% | 0.8% |
| Depressive disorder | 2,533 | 7.8% | 43.6% | 40.2% | 8.5% | 8.4% | 12.4% | 16.9% | 7.6% | - | 4.1% | 4.3% | 7.1% | 1.1% | 2.0% | 0.4% | 1.0% | 0.6% | 5.3% | 2.2% | 0.3% |
| Neurodevelopmental disorder: other | 790 | 2.4% | 51.9% | 21.8% | 10.1% | 15.7% | 2.8% | 4.6% | 3.7% | 2.9% | - | 0.5% | 17.5% | 3.9% | 0.6% | 1.0% | 0.1% | 0.1% | 0.5% | 0.5% | 0.8% |
| Feeding and eating disorder | 1,332 | 4.1% | 68.1% | 14.5% | 5.8% | 4.3% | 3.6% | 7.9% | 1.7% | 11.9% | 1.4% | - | 2.6% | 2.6% | 0.5% | 0.7% | 0.1% | 0.1% | 1.6% | 0.1% | 0.1% |
| Neurodevelopmental disorder: intellectual disabilities | 587 | 1.8% | 51.6% | 27.4% | 13.3% | 16.7% | 8.5% | 5.3% | 14.8% | 2.9% | 3.7% | 0.0% | - | 1.5% | 0.2% | 0.2% | 0.2% | 1.2% | 0.0% | 1.0% | 1.0% |
| Obsessive-compulsive and related disorders | 653 | 2.0% | 53.8% | 20.8% | 12.9% | 7.8% | 4.0% | 13.5% | 1.5% | 7.5% | 5.8% | 4.0% | 7.7% | - | 1.2% | 0.5% | 0.2% | 0.2% | 2.0% | 0.2% | 0.2% |
| Somatic symptom and related disorders | 597 | 1.8% | 60.8% | 21.9% | 5.9% | 6.4% | 5.7% | 11.6% | 1.5% | 10.6% | 5.9% | 1.3% | 5.2% | 0.8% | - | 0.3% | 0.2% | 0.3% | 2.0% | 0.0% | 0.8% |
| Other mental disorders | 155 | 0.5% | 64.5% | 16.1% | 1.9% | 2.6% | 1.9% | 3.2% | 3.2% | 2.6% | 2.6% | 1.3% | 16.1% | 0.0% | 3.2% | - | 0.0% | 0.0% | 0.0% | 1.3% | 0.6% |
| Gender dysphoria | 203 | 0.6% | 72.9% | 47.3% | 4.4% | 3.9% | 4.9% | 4.4% | 2.0% | 11.8% | 4.4% | 0.5% | 1.5% | 0.5% | 0.5% | 1.0% | - | 0.5% | 1.5% | 0.0% | 0.0% |
| Schizophrenia spectrum and other psychotic disorders | 208 | 0.6% | 36.5% | 18.8% | 21.2% | 7.2% | 9.1% | 5.8% | 7.2% | 5.8% | 4.3% | 1.0% | 30.3% | 2.4% | 1.0% | 1.0% | 0.5% | - | 2.4% | 7.7% | 0.0% |
| Personality disorder | 197 | 0.6% | 22.8% | 52.3% | 13.2% | 17.3% | 17.3% | 20.3% | 13.7% | 29.4% | 2.5% | 13.2% | 6.6% | 1.0% | 3.0% | 0.0% | 0.5% | 1.0% | - | 3.6% | 0.0% |
| Substance-related and addictive disorders | 121 | 0.4% | 38.8% | 42.1% | 9.1% | 13.2% | 8.3% | 5.0% | 27.3% | 7.4% | 0.0% | 0.8% | 14.0% | 0.0% | 1.7% | 0.8% | 0.0% | 1.7% | 4.1% | - | 0.8% |
| Elimination disorder | 24 | 0.1% | - | - | - | - | - | - | - | - | - | - | - | - | - | - | - | - | - | - | - |
