## Supplemental table 8 for "Patterns of mental disorders in a nationwide child psychiatric sample (*N*=67,815): A DREAMS study"

Table S 8. Primary and five year comorbid DSM-5 diagnoses in males aged 0 through 12 years at time of primary diagnosis (*N* = 11,223).

| | Total | | | | Comorbid diagnosis | | | | | | | | | | | | | | | | |
|---|---|---|---|---|---|---|---|---|---|---|---|---|---|---|---|---|---|---|---|---|---|
| | | | No comorbid diagnosis | V-code | Neurodevelopmental disorder: ASD | Neurodevelopmental disorder: ADHD | Trauma and stressor-related disorders | Anxiety disorders | Disruptive. impulse-control and conduct disorders | Depressive disorder | Neurodevelopmental disorder: other | Feeding and eating disorder | Neurodevelopmental disorder: intellectual disabilities | Obsessive-compulsive and related disorders | Somatic symptom and related disorders | Other mental disorders | Gender dysphoria | Schizophrenia spectrum and other psychotic disorders | Personality disorder | Substance-related and addictive disorders | Elimination disorder |
| **Primary diagnosis** | n | % | % | % | % | % | % | % | % | % | % | % | % | % | % | % | % | % | % | % | % |
| Neurodevelopmental disorder: ASD | 4,896 | 43.6% | 53.5% | 23.1% | - | 22.0% | 1.8% | 2.2% | 3.4% | 0.7% | 10.5% | 0.7% | 15.8% | 0.4% | 0.1% | 0.2% | 0.0% | 0.0% | 0.0% | 0.0% | 2.1% |
| Neurodevelopmental disorder: ADHD | 3,518 | 31.3% | 60.1% | 30.2% | 10.1% | - | 2.6% | 2.3% | 9.4% | 0.7% | 11.0% | 0.1% | 12.4% | 0.3% | 0.1% | 0.4% | 0.0% | 0.0% | 0.0% | 0.0% | 1.4% |
| Trauma and stressor-related disorders | 698 | 6.2% | 61.6% | 34.1% | 4.2% | 14.0% | - | 3.0% | 8.2% | 0.9% | 4.4% | 0.4% | 12.5% | 0.1% | 0.0% | 0.3% | 0.0% | 0.0% | 0.0% | 0.1% | 2.7% |
| Anxiety disorders | 469 | 4.2% | 69.9% | 24.3% | 3.6% | 8.3% | 3.8% | - | 2.6% | 1.7% | 6.8% | 0.6% | 7.2% | 0.9% | 1.1% | 0.2% | 0.6% | 0.0% | 0.2% | 0.0% | 2.1% |
| Disruptive, impulse-control and conduct disorders | 534 | 4.8% | 62.4% | 37.5% | 5.4% | 15.7% | 4.1% | 3.7% | - | 0.6% | 5.8% | 0.4% | 10.3% | 0.0% | 0.0% | 0.6% | 0.2% | 0.0% | 0.0% | 0.0% | 1.5% |
| Depressive disorder | 64 | 0.6% | 50.0% | 48.4% | 10.9% | 18.8% | 4.7% | 10.9% | 4.7% | - | 7.8% | 1.6% | 6.3% | 0.0% | 0.0% | 0.0% | 0.0% | 0.0% | 0.0% | 0.0% | 3.1% |
| Neurodevelopmental disorder: other | 616 | 5.5% | 64.9% | 27.3% | 7.8% | 8.4% | 0.6% | 2.9% | 3.2% | 0.5% | - | 0.5% | 14.4% | 0.8% | 0.0% | 1.0% | 0.0% | 0.0% | 0.0% | 0.0% | 1.1% |
| Feeding and eating disorder | 57 | 0.5% | 77.2% | 8.8% | 7.0% | 7.0% | 1.8% | 1.8% | 0.0% | 0.0% | 3.5% | - | 3.5% | 0.0% | 0.0% | 0.0% | 0.0% | 0.0% | 0.0% | 0.0% | 3.5% |
| Neurodevelopmental disorder: intellectual disabilities | 177 | 1.6% | 67.2% | 20.9% | 14.1% | 10.7% | 1.7% | 0.6% | 3.4% | 0.0% | 4.5% | 0.6% | - | 0.0% | 0.0% | 0.0% | 0.0% | 0.0% | 0.0% | 0.0% | 1.1% |
| Obsessive-compulsive and related disorders | 44 | 0.4% | - | - | - | - | - | - | - | - | - | - | - | - | - | - | - | - | - | - | - |
| Somatic symptom and related disorders | 18 | 0.2% | - | - | - | - | - | - | - | - | - | - | - | - | - | - | - | - | - | - | - |
| Other mental disorders | 61 | 0.5% | 75.4% | 13.1% | 3.3% | 1.6% | 3.3% | 1.6% | 4.9% | 0.0% | 1.6% | 0.0% | 16.4% | 0.0% | 0.0% | - | 0.0% | 0.0% | 0.0% | 0.0% | 0.0% |
| Gender dysphoria | 8 | 0.1% | - | - | - | - | - | - | - | - | - | - | - | - | - | - | - | - | - | - | - |
| Schizophrenia spectrum and other psychotic disorders | 1 | 0.0% | - | - | - | - | - | - | - | - | - | - | - | - | - | - | - | - | - | - | - |
| Personality disorder | 3 | 0.0% | - | - | - | - | - | - | - | - | - | - | - | - | - | - | - | - | - | - | - |
| Substance-related and addictive disorders | 1 | 0.0% | - | - | - | - | - | - | - | - | - | - | - | - | - | - | - | - | - | - | - |
| Elimination disorder | 29 | 0.3% | - | - | - | - | - | - | - | - | - | - | - | - | - | - | - | - | - | - | - |
