## Supplemental table 9 for "Patterns of mental disorders in a nationwide child psychiatric sample (*N*=67,815): A DREAMS study"

Table S 9. Primary and five year comorbid DSM-5 diagnoses in males aged 13 through 23 years at time of primary diagnosis (*N* = 18,432).

| | Total | | | | Comorbid diagnosis | | | | | | | | | | | | | | | | |
|---|---|---|---|---|---|---|---|---|---|---|---|---|---|---|---|---|---|---|---|---|---|
| | | | No comorbid diagnosis | V-code | Neurodevelopmental disorder: ASD | Neurodevelopmental disorder: ADHD | Trauma and stressor-related disorders | Anxiety disorders | Disruptive, impulse-control and conduct disorders | Depressive disorder | Neurodevelopmental disorder: other | Feeding and eating disorder | Neurodevelopmental disorder: intellectual disabilities | Obsessive-compulsive and related disorders | Somatic symptom and related disorders | Other mental disorders | Gender dysphoria | Schizophrenia spectrum and other psychotic disorders | Personality disorder | Substance-related and addictive disorders | Elimination disorder |
| **Primary diagnosis** | n | % | % | % | % | % | % | % | % | % | % | % | % | % | % | % | % | % | % | % | % |
| Neurodevelopmental disorder: ASD | 7,260 | 39.4% | 44.5% | 24.5% | - | 23.4% | 2.9% | 4.5% | 5.5% | 4.8% | 10.9% | 0.6% | 18.7% | 1.3% | 0.6% | 0.3% | 0.2% | 0.6% | 0.4% | 0.8% | 1.1% |
| Neurodevelopmental disorder: ADHD | 5,037 | 27.3% | 51.3% | 23.0% | 9.1% | - | 4.5% | 3.4% | 14.4% | 2.8% | 10.7% | 0.2% | 16.3% | 0.5% | 0.2% | 0.1% | 0.1% | 0.1% | 0.4% | 1.7% | 1.3% |
| Trauma and stressor-related disorders | 1,138 | 6.2% | 44.7% | 31.4% | 6.5% | 18.1% | - | 5.0% | 15.0% | 8.0% | 3.3% | 0.2% | 20.0% | 0.5% | 0.1% | 0.4% | 0.1% | 0.4% | 1.1% | 3.9% | 1.5% |
| Anxiety disorders | 933 | 5.1% | 56.5% | 26.6% | 8.9% | 10.8% | 5.5% | - | 2.4% | 8.3% | 6.6% | 1.0% | 11.0% | 1.8% | 1.7% | 0.1% | 0.2% | 0.3% | 2.1% | 1.8% | 0.3% |
| Disruptive, impulse-control and conduct disorders | 1,346 | 7.3% | 44.3% | 37.6% | 7.3% | 20.0% | 8.2% | 2.5% | - | 3.5% | 5.3% | 0.3% | 21.7% | 0.1% | 0.1% | 0.3% | 0.1% | 0.3% | 1.8% | 7.0% | 1.0% |
| Depressive disorder | 758 | 4.1% | 44.3% | 39.7% | 14.6% | 11.1% | 8.8% | 14.4% | 6.3% | - | 7.1% | 0.7% | 8.3% | 1.3% | 1.8% | 0.1% | 1.3% | 0.7% | 4.1% | 3.6% | 0.4% |
| Neurodevelopmental disorder: other | 549 | 3.0% | 51.5% | 20.4% | 10.0% | 17.5% | 2.6% | 4.4% | 4.0% | 2.4% | - | 0.2% | 18.2% | 3.3% | 0.5% | 0.7% | 0.0% | 0.2% | 0.2% | 0.5% | 0.9% |
| Feeding and eating disorder | 116 | 0.6% | 65.5% | 12.9% | 12.9% | 6.9% | 1.7% | 6.9% | 0.0% | 6.9% | 0.9% | - | 5.2% | 5.2% | 0.9% | 0.9% | 0.0% | 0.0% | 0.0% | 0.0% | 0.0% |
| Neurodevelopmental disorder: intellectual disabilities | 384 | 2.1% | 46.1% | 27.1% | 16.7% | 19.3% | 8.6% | 4.2% | 17.7% | 2.9% | 4.4% | 0.0% | - | 1.0% | 0.0% | 0.0% | 0.3% | 0.8% | 0.0% | 1.3% | 1.3% |
| Obsessive-compulsive and related disorders | 232 | 1.3% | 51.3% | 15.9% | 17.7% | 11.6% | 4.7% | 11.2% | 1.3% | 6.0% | 9.1% | 1.7% | 5.6% | - | 0.9% | 0.0% | 0.4% | 0.4% | 0.9% | 0.4% | 0.4% |
| Somatic symptom and related disorders | 189 | 1.0% | 53.4% | 22.2% | 11.6% | 8.5% | 6.3% | 12.2% | 2.1% | 7.9% | 9.0% | 0.5% | 4.8% | 2.1% | - | 0.0% | 0.0% | 0.0% | 2.1% | 0.0% | 2.1% |
| Other mental disorders | 81 | 0.4% | 72.8% | 14.8% | 1.2% | 3.7% | 1.2% | 1.2% | 3.7% | 1.2% | 4.9% | 1.2% | 11.1% | 0.0% | 1.2% | - | 0.0% | 0.0% | 0.0% | 2.5% | 1.2% |
| Gender dysphoria | 89 | 0.5% | 75.3% | 51.7% | 4.5% | 3.4% | 4.5% | 2.2% | 1.1% | 12.4% | 5.6% | 1.1% | 1.1% | 1.1% | 1.1% | 1.1% | - | 1.1% | 1.1% | 0.0% | 0.0% |
| Schizophrenia spectrum and other psychotic disorders | 121 | 0.7% | 33.9% | 16.5% | 24.8% | 9.1% | 10.7% | 4.1% | 9.1% | 5.0% | 4.1% | 0.0% | 31.4% | 2.5% | 1.7% | 0.8% | 0.8% | - | 0.8% | 10.7% | 0.0% |
| Personality disorder | 40 | 0.2% | - | - | - | - | - | - | - | - | - | - | - | - | - | - | - | - | - | - | - |
| Substance-related and addictive disorders | 91 | 0.5% | 37.4% | 42.9% | 11.0% | 15.4% | 5.5% | 6.6% | 24.2% | 8.8% | 0.0% | 1.1% | 12.1% | 0.0% | 2.2% | 0.0% | 0.0% | 2.2% | 3.3% | - | 1.1% |
| Elimination disorder | 16 | 0.1% | - | - | - | - | - | - | - | - | - | - | - | - | - | - | - | - | - | - | - |
