## Supplemental table 10 for "Patterns of mental disorders in a nationwide child psychiatric sample (*N*=67,815): A DREAMS study"

Table S 10. Primary and five year comorbid DSM-5 diagnoses in females aged 0 through 12 years at time of primary diagnosis ($N$ = 4,418).

| | Total | | Comorbid diagnosis | | | | | | | | | | | | | | | | | | |
|---|---|---|---|---|---|---|---|---|---|---|---|---|---|---|---|---|---|---|---|---|---|
| | | | No comorbid diagnosis | V-code | Neurodevelopmental disorder: ASD | Neurodevelopmental disorder: ADHD | Trauma and stressor-related disorders | Anxiety disorders | Disruptive, impulse-control and conduct disorders | Depressive disorder | Neurodevelopmental disorder: other | Feeding and eating disorder | Neurodevelopmental disorder: intellectual disabilities | Obsessive-compulsive and related disorders | Somatic symptom and related disorders | Other mental disorders | Gender dysphoria | Schizophrenia spectrum and other psychotic disorders | Personality disorder | Substance-related and addictive disorders | Elimination disorder |
| **Primary diagnosis** | n | % | % | % | % | % | % | % | % | % | % | % | % | % | % | % | % | % | % | % | % |
| Neurodevelopmental disorder: ASD | 1,314 | 29.7% | 57.3% | 27.5% | - | 15.9% | 1.0% | 3.9% | 2.2% | 1.3% | 7.8% | 1.2% | 15.3% | 0.9% | 0.5% | 0.2% | 0.2% | 0.0% | 0.2% | 0.0% | 1.4% |
| Neurodevelopmental disorder: ADHD | 1,165 | 26.4% | 64.4% | 31.8% | 7.5% | - | 3.1% | 3.5% | 6.1% | 0.5% | 9.1% | 0.6% | 12.3% | 0.3% | 0.3% | 0.6% | 0.1% | 0.0% | 0.2% | 0.0% | 1.6% |
| Trauma and stressor-related disorders | 525 | 11.9% | 70.1% | 37.5% | 2.7% | 8.4% | - | 2.7% | 2.9% | 1.3% | 2.3% | 0.6% | 13.7% | 0.0% | 0.0% | 0.2% | 0.2% | 0.0% | 0.0% | 0.0% | 0.8% |
| Anxiety disorders | 597 | 13.5% | 78.7% | 22.6% | 3.5% | 4.5% | 2.3% | - | 1.5% | 1.2% | 5.2% | 0.0% | 4.9% | 1.3% | 1.5% | 0.3% | 0.2% | 0.0% | 0.3% | 0.0% | 0.2% |
| Disruptive, impulse-control and conduct disorders | 183 | 4.1% | 68.3% | 44.3% | 2.2% | 6.6% | 4.9% | 3.3% | - | 0.0% | 6.0% | 0.0% | 12.0% | 0.0% | 0.0% | 0.5% | 0.0% | 0.0% | 0.0% | 0.0% | 0.5% |
| Depressive disorder | 65 | 1.5% | 50.8% | 61.5% | 7.7% | 10.8% | 7.7% | 9.2% | 7.7% | - | 4.6% | 3.1% | 6.2% | 0.0% | 0.0% | 1.5% | 0.0% | 0.0% | 3.1% | 0.0% | 0.0% |
| Neurodevelopmental disorder: other | 243 | 5.5% | 65.8% | 22.2% | 3.7% | 10.3% | 1.2% | 3.3% | 2.1% | 0.4% | - | 0.8% | 18.5% | 0.4% | 0.4% | 0.4% | 0.0% | 0.0% | 0.4% | 0.0% | 0.0% |
| Feeding and eating disorder | 63 | 1.4% | 68.3% | 15.9% | 4.8% | 0.0% | 4.8% | 6.3% | 6.3% | 3.2% | 3.2% | - | 4.8% | 1.6% | 1.6% | 0.0% | 0.0% | 0.0% | 1.6% | 0.0% | 0.0% |
| Neurodevelopmental disorder: intellectual disabilities | 98 | 2.2% | 77.6% | 24.5% | 12.2% | 7.1% | 4.1% | 0.0% | 3.1% | 0.0% | 1.0% | 0.0% | - | 0.0% | 0.0% | 0.0% | 0.0% | 0.0% | 0.0% | 0.0% | 0.0% |
| Obsessive-compulsive and related disorders | 58 | 1.3% | 60.3% | 20.7% | 10.3% | 5.2% | 3.4% | 25.9% | 0.0% | 0.0% | 3.4% | 0.0% | 1.7% | - | 0.0% | 0.0% | 0.0% | 0.0% | 1.7% | 0.0% | 0.0% |
| Somatic symptom and related disorders | 22 | 0.5% | - | - | - | - | - | - | - | - | - | - | - | - | - | - | - | - | - | 0.0% | 0.0% |
| Other mental disorders | 41 | 0.9% | - | - | - | - | - | - | - | - | - | - | - | - | - | - | - | - | - | 0.0% | 0.0% |
| Gender dysphoria | 14 | 0.3% | - | - | - | - | - | - | - | - | - | - | - | - | - | - | - | - | - | 0.0% | 0.0% |
| Schizophrenia spectrum and other psychotic disorders | 0 | 0.0% | - | - | - | - | - | - | - | - | - | - | - | - | - | - | - | - | - | - | - |
| Personality disorder | 3 | 0.1% | - | - | - | - | - | - | - | - | - | - | - | - | - | - | - | - | - | 0.0% | 0.0% |
| Substance-related and addictive disorders | 0 | 0.0% | - | - | - | - | - | - | - | - | - | - | - | - | - | - | - | - | - | - | - |
| Elimination disorder | 14 | 0.3% | - | - | - | - | - | - | - | - | - | - | - | - | - | - | - | - | - | 0.0% | - |
