## Supplemental table 11 for "Patterns of mental disorders in a nationwide child psychiatric sample (*N*=67,815): A DREAMS study"

Table S 11. Primary and five year comorbid DSM-5 diagnoses in females aged 13 through 23 years at time of primary diagnosis (*N* = 14,076).

| | Total | | No comorbid diagnosis | V-code | Comorbid diagnosis | | | | | | | | | | | | | | | | |
|---|---|---|---|---|---|---|---|---|---|---|---|---|---|---|---|---|---|---|---|---|---|
| | | | | | Neurodevelopmental disorder: ASD | Neurodevelopmental disorder: ADHD | Trauma and stressor-related disorders | Anxiety disorders | Disruptive, impulse-control and conduct disorders | Depressive disorder | Neurodevelopmental disorder: other | Feeding and eating disorder | Neurodevelopmental disorder: intellectual disabilities | Obsessive-compulsive and related disorders | Somatic symptom and related disorders | Other mental disorders | Gender dysphoria | Schizophrenia spectrum and other psychotic disorders | Personality disorder | Substance-related and addictive disorders | Elimination disorder |
| **Primary diagnosis** | n | % | % | % | % | % | % | % | % | % | % | % | % | % | % | % | % | % | % | % | % |
| Neurodevelopmental disorder: ASD | 2,982 | 21.2% | 42.3% | 29.8% | - | 13.8% | 5.1% | 10.0% | 3.7% | 12.1% | 6.8% | 4.0% | 17.2% | 2.8% | 1.6% | 0.4% | 0.5% | 0.6% | 1.4% | 0.3% | 0.8% |
| Neurodevelopmental disorder: ADHD | 1,978 | 14.1% | 53.2% | 28.2% | 4.7% | - | 6.7% | 6.3% | 10.9% | 6.1% | 8.0% | 1.1% | 13.3% | 0.7% | 0.9% | 0.3% | 0.1% | 0.2% | 2.2% | 0.7% | 0.7% |
| Trauma and stressor-related disorders | 1,804 | 12.8% | 48.6% | 33.8% | 3.4% | 9.5% | - | 7.2% | 8.3% | 13.7% | 2.7% | 3.3% | 15.0% | 1.0% | 1.9% | 0.3% | 0.3% | 0.6% | 3.8% | 2.4% | 0.3% |
| Anxiety disorders | 1,721 | 12.2% | 53.9% | 26.4% | 5.8% | 5.8% | 5.9% | - | 2.8% | 16.7% | 4.1% | 3.0% | 9.4% | 2.6% | 2.7% | 0.1% | 0.3% | 0.2% | 3.4% | 0.5% | 0.2% |
| Disruptive, impulse-control and conduct disorders | 811 | 5.8% | 46.1% | 48.7% | 4.7% | 12.5% | 11.6% | 6.5% | - | 12.8% | 4.1% | 3.0% | 16.4% | 0.7% | 0.4% | 0.9% | 0.1% | 0.4% | 2.2% | 4.7% | 0.5% |
| Depressive disorder | 1,775 | 12.6% | 43.3% | 40.4% | 5.9% | 7.3% | 13.9% | 18.0% | 8.2% | - | 2.8% | 5.8% | 6.5% | 1.0% | 2.1% | 0.5% | 0.9% | 0.5% | 5.9% | 1.6% | 0.3% |
| Neurodevelopmental disorder: other | 241 | 1.7% | 52.7% | 24.9% | 10.4% | 11.6% | 3.3% | 5.0% | 2.9% | 4.1% | - | 1.2% | 15.8% | 5.4% | 0.8% | 1.7% | 0.4% | 0.0% | 1.2% | 0.4% | 0.4% |
| Feeding and eating disorder | 1,216 | 8.6% | 68.3% | 14.6% | 5.1% | 4.0% | 3.8% | 8.0% | 1.9% | 12.4% | 1.4% | - | 2.3% | 2.3% | 0.5% | 0.7% | 0.1% | 0.1% | 1.7% | 0.1% | 0.1% |
| Neurodevelopmental disorder: intellectual disabilities | 203 | 1.4% | 62.1% | 28.1% | 6.9% | 11.8% | 8.4% | 7.4% | 9.4% | 3.0% | 2.5% | 0.0% | - | 2.5% | 0.5% | 0.5% | 0.0% | 2.0% | 0.0% | 0.5% | 0.5% |
| Obsessive-compulsive and related disorders | 421 | 3.0% | 55.1% | 23.5% | 10.2% | 5.7% | 3.6% | 14.7% | 1.7% | 8.3% | 4.0% | 5.2% | 8.8% | - | 1.4% | 0.7% | 0.0% | 0.0% | 2.6% | 0.0% | 0.0% |
| Somatic symptom and related disorders | 408 | 2.9% | 64.2% | 21.8% | 3.2% | 5.4% | 5.4% | 11.3% | 1.2% | 11.8% | 4.4% | 1.7% | 5.4% | 0.2% | - | 0.5% | 0.2% | 0.5% | 2.0% | 0.0% | 0.2% |
| Other mental disorders | 74 | 0.5% | 55.4% | 17.6% | 2.7% | 1.4% | 2.7% | 5.4% | 2.7% | 4.1% | 0.0% | 1.4% | 21.6% | 0.0% | 5.4% | - | 0.0% | 0.0% | 0.0% | 0.0% | 0.0% |
| Gender dysphoria | 114 | 0.8% | 71.1% | 43.9% | 4.4% | 4.4% | 5.3% | 6.1% | 2.6% | 11.4% | 3.5% | 0.0% | 1.8% | 0.0% | 0.0% | 0.9% | - | 0.0% | 1.8% | 0.0% | 0.0% |
| Schizophrenia spectrum and other psychotic disorders | 87 | 0.6% | 40.2% | 21.8% | 16.1% | 4.6% | 6.9% | 8.0% | 4.6% | 6.9% | 4.6% | 2.3% | 28.7% | 2.3% | 0.0% | 1.1% | 0.0% | - | 4.6% | 3.4% | 0.0% |
| Personality disorder | 157 | 1.1% | 20.4% | 56.1% | 15.9% | 16.6% | 17.2% | 22.9% | 11.5% | 31.8% | 3.2% | 16.6% | 5.1% | 0.6% | 3.8% | 0.0% | 0.6% | 0.6% | - | 1.9% | 0.0% |
| Substance-related and addictive disorders | 30 | 0.2% | - | - | - | - | - | - | - | - | - | - | - | - | - | - | - | - | - | - | - |
| Elimination disorder | 8 | 0.1% | - | - | - | - | - | - | - | - | - | - | - | - | - | - | - | - | - | - | - |
