## Supplementary figures and images for "Patterns of mental disorders in a nationwide child psychiatric sample (*N*=67,815): A DREAMS study"

### Supplemental figure 1

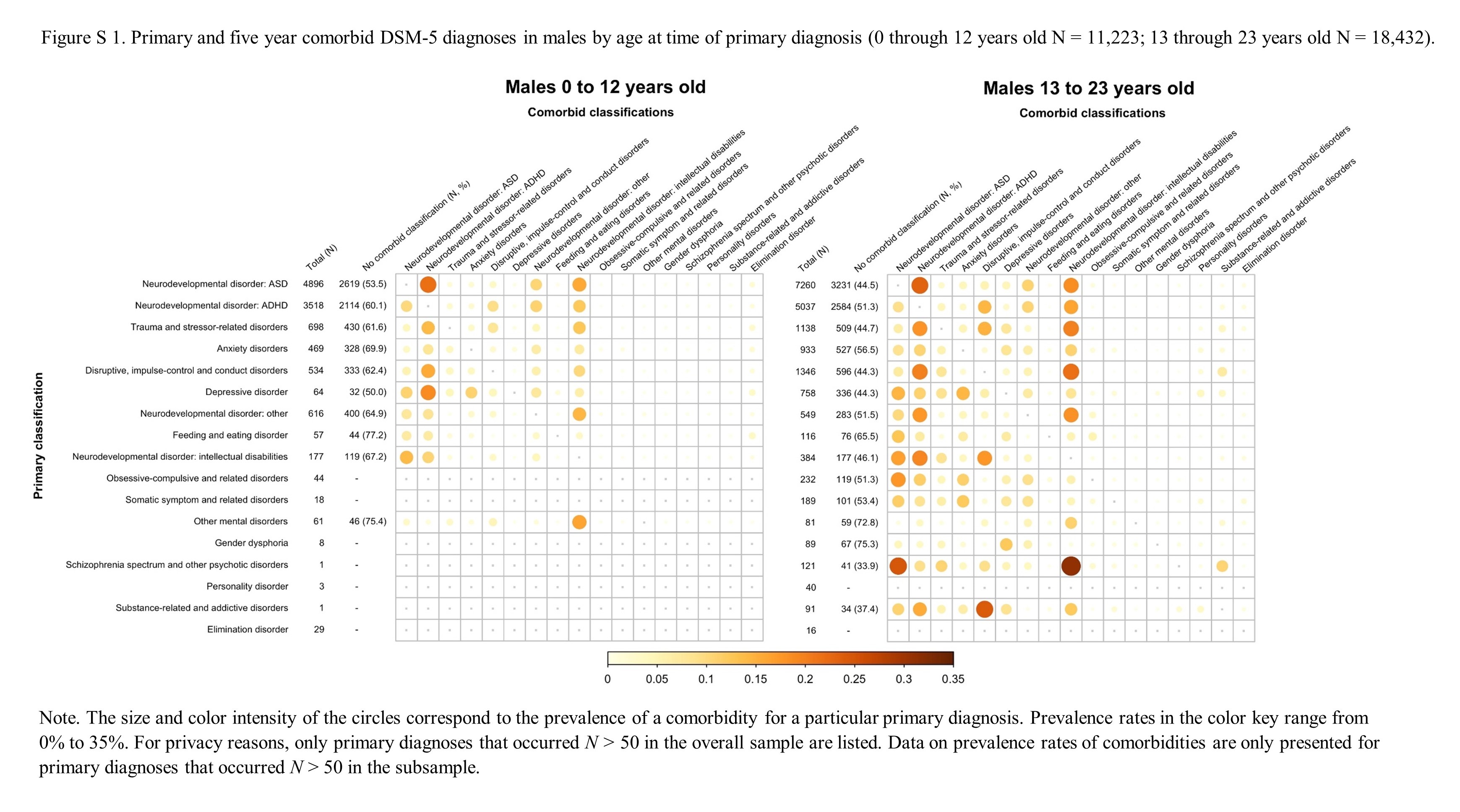

### Supplemental figure 2

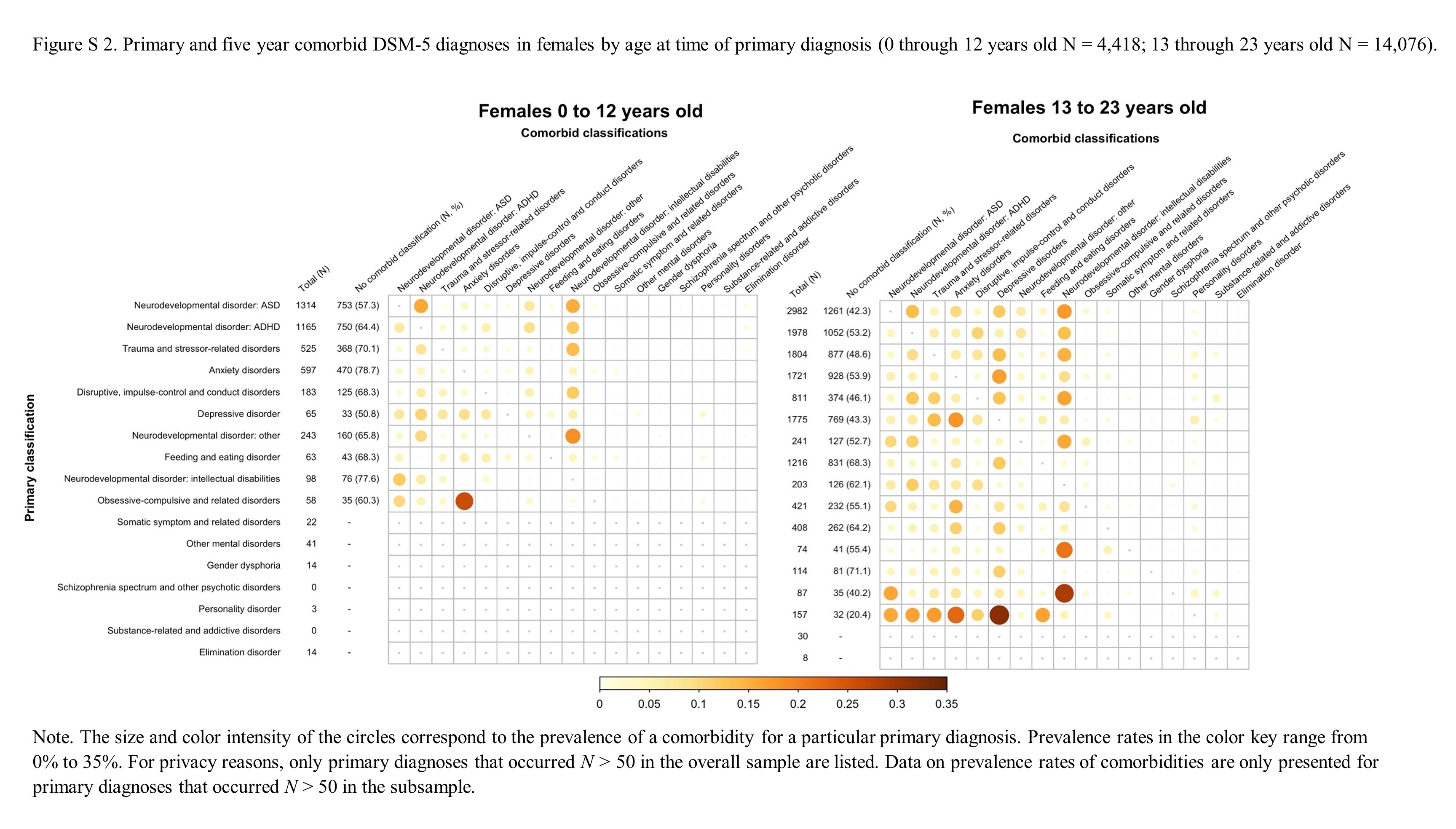
